# AnFiSA: An open-source computational platform for the analysis of sequencing data for rare genetic disease

**DOI:** 10.1101/2021.09.26.21263358

**Authors:** M.A. Bouzinier, D. Etin, S.I. Trifonov, V.N. Evdokimova, V. Ulitin, J. Shen, A. Kokorev, A. A. Ghazani, Y. Chekaluk, Z. Albertyn, A. Giersch, C.C. Morton, F. Abraamyan, P.K. Bendapudi, S. Sunyaev, Undiagnosed Diseases Network, Brigham Genomic Medicine, SEQuencing a Baby for an Optimal Outcome, Quantori, J.B. Krier

**Affiliations:** Division of Genetics, Department of Medicine, Brigham and Women’s Hospital, Harvard Medical School, Boston, MA, USA; Forome Association, Boston, MA, USA; Oracle Corporation, USA; SBCS Scientific Biomedical Consulting Services, London, UK; Department of Pathology, Brigham and Women’s Hospital, Harvard Medical School, Boston, MA, USA; ITMO University, St. Petersburg, Russian Federation; Division of Hemostasis and Thrombosis, Beth Israel Deaconess Medical Center, Boston, MA, USA; Division of Hematology and Blood Transfusion Service, Massachusetts General Hospital, Boston, MA, USA; Harvard Medical School, Boston, MA, USA; Undiagnosed Diseases Network, NIH; Brigham Genomic Medicine, Brigham and Women’s Hospital, Boston, MA, USA; Quantori, Cambridge, MA, USA; Department of Obstetrics and Gynecology, Brigham and Women’s Hospital, Harvard Medical School, Boston, MA, USA; Broad Institute of MIT and Harvard, Cambridge, MA, USA; Manchester Centre for Audiology and Deafness (ManCAD), School of Health Sciences, University of Manchester, UK

## Abstract

Despite genomic sequencing rapidly transforming from being a bench-side tool to a routine procedure in a hospital, there is a noticeable lack of genomic analysis software that supports both clinical and research workflows as well as crowdsourcing. Furthermore, most existing software packages are not forward-compatible in regards to supporting ever-changing diagnostic rules adopted by the genetics community. Regular updates of genomics databases pose challenges for reproducible and traceable automated genetic diagnostics tools. Lastly, most of the software tools score low on explainability amongst clinicians.

We have created a fully open-source variant curation tool, *AnFiSA,* with the intention to invite and accept contributions from clinicians, researchers and professional software developers. The design of *AnFiSA* addresses the aforementioned issues via the following architectural principles: using a multidimensional database management system (DBMS) for genomic data to address reproducibility, curated decision trees adaptable to changing clinical rules, and a crowdsourcing-friendly interface to address difficult-to-diagnose cases. We discuss how we have chosen our technology stack and describe the design and implementation of the software. Finally, we show in detail how selected workflows can be implemented using the current version of *AnFiSA* by a medical geneticist.

All software is available at https://github.com/ForomePlatform under the Apache 2.0 license.

The public demo instance with the public data sets can be accessed via https://github.com/ForomePlatform/AnFiSA#public-demo

## Introduction

Genomic sequencing has proven to be an effective research and diagnostic tool for rare monogenic diseases [1], [2]. NGS-based assays, first implemented over a decade ago, have now become the prevalent technology in clinical genomics [3]–[5]. NGS is a high throughput technology which can generate an extremely large amount of genetic sequence variants in one assay, which is both its greatest advantage and presents a significant challenge for those who wish to implement NGS-based workflows.

We present a software platform to facilitate NGS based research and diagnostic workflows in germline genomics of rare Mendelian diseases: Annotation and Filtration for Sequencing Analysis (*AnFiSA)*. The goal of the project is to enable the seamless transformation of research workflows into novel clinical protocols by building a notebook-like environment [6] for the design and iteration of clinical analysis. Moreover, the platform has been developed to invite and accept contributions from clinicians, researchers and professional software developers.

### Defining the challenge: Genomic sequencing analysis for monogenic disease diagnosis

The processing and interpretation of NGS data requires multiple steps and more computational power and storage than almost all other tasks in the biomedical sciences. The following overview illustrates the challenges to be addressed by a genomics analysis and interpretation platform such as *AnFiSA*.

When working with unknown Mendelian diseases, it is common to analyze exome and genome sequences. A number of approaches are used to architect upstream genomics analysis pipelines [7], [8]. Upstream pipelines intake and process raw BAM or FASTQ files consuming 100s of GBs of data, and include alignment, recalibration and deduplication, variant calling, and storage of large temporary files. This workflow is well defined, based on GATK Best Practices[9]–[11]. Downstream genomic sequencing analysis is less standardized and does not have a dominating set of tools and approaches. In downstream analysis, the primary input is the Variant Call Format file (VCF), and a sequence of steps or a pipeline which requires highly multidimensional and multivalued annotation and subsequent filtration of millions of variants. Preparation of multidimensional and multivalued data for efficient annotation, filtration, and storage requires a flexible data model and efficient indexing mechanisms, thus presenting a design challenge. A separate challenge is the visualization of the data that can be used for curation and interpretation of the NGS output in clinical and scientific research. In comparison to upstream analysis where most of the bottlenecks are computational and thus can be solved with High Performance Computing (HPC), the downstream analysis requires development and a combination of fit-for-purpose software tools.

Initially, clinical interpretation of sequence variants was done manually, without structured protocols. During and after the development of clinical NGS-based assays, multiple laboratories independently established algorithms, tools and approaches for variant analysis and reporting. In 2015, a joint consensus recommendation of the ACMG and the Association for Molecular Pathology (AMP) for variant interpretation was published [12]. ACMG/AMP variant interpretation guidelines recommended using information from the population, disease-specific and sequence databases, and *in-silico* computational predictive programs for variant curation in addition to information obtained from peer-reviewed publications such as genotype-phenotype segregation and functional data. They simplified and made consistent variant interpretation between and within laboratories. This resulted in an increase and stabilization of diagnostic rates for clinical tests which utilized the NGS platform [13]. The number of variants of uncertain significance (VUS) were also decreased [13], where the VUS detection rate was linked to the quality and efficacy of variant curation [13], [14]. Alternative classification approaches are also being explored [15].

Though a number of computational tools emerged for variant interpretation, the typical research or diagnostic problem consists of identifying the pathogenic mutation amongst the myriad of allelic variants in the patient’s genome. In both clinical and research contexts, candidate pathogenic mutations are prioritized based on features such as population allele frequency and predicted impact on molecular function. Prioritization rules and workflows vary depending on disease type, diagnostic or research setting, mode of inheritance and number of study subjects (an individual proband, a nuclear family, a larger pedigree or a cohort of unrelated patients). Conceptually, this process is not different from data analysis workflows in other fields, including fields outside of life sciences, where data are represented by multiple features that are the basis for an inference or predictive model.

Despite gradual improvement, there is still lack of codified algorithms for variant curation to answer many relevant questions in human genetics. We see a division in development of the software tools for standardized clinical tasks where the codified algorithms can answer limited questions, and the research-focused tools, that are less automated but allow customized queries, thus compromising on repeatability, reproducibility and traceability.

When a monogenic etiology for a disease is suspected, evaluation of patients with medically complicated conditions requires both thorough evaluation of a patient’s clinical features as well as intensive interrogation of relevant scientific reports, which is impossible without having a transparent and traceable work process. Reproducibility [16] and traceability are an integral part of laboratory quality management processes governed by Good Clinical Practice guidelines [17] or Clinical Laboratory Improvement Amendments (CLIA) requirements [18]. Many proprietary commercial curation tools use opaque machine learning and artificial intelligence models to deliver the treating clinician a conclusion while often sacrificing explainability and therefore traceability of the results [19]–[21]. For example, a high-throughput pipeline built by Clark et al [22] focuses on the minimal user interaction and shortening the time to delivering a provisional finding. Others [23] take explainability quite seriously, nevertheless giving clinician little room for adjudication. The notion of minimal user interaction might be disputed given results shown by Basel-Salmon et al [24].

Taking all the above in consideration, it would be fair to say, that the clinical community working with complex clinical cases such as undiagnosed diseases with unknown genetic etiology does not have an optimal set of supporting tools.

### Development landscape and design principles

*AnFiSA* was designed to be a specialized data warehouse for complex genomic data analysis, such as whole genome sequencing for patients with rare diseases. The lack of robust, open-source software to address the challenges clinical genomic data present may seem surprising given that, outside the fields of genetics and genomics, there is a thriving community that continuously produces open-source software for data analysis, and cumulatively covers virtually all aspects of data manipulation and exploration. *Apache Software Foundation* (ASF), for example, alone hosts approximately 50 projects dedicated to data analysis, but none of them are dedicated to genomic data. In contrast, most genomic analysis tools are either a) proprietary and require significant financial costs or data sharing obligations; or b) academically developed and often under-documented, unmaintained, and/or deprecated. The small number of open-source and well-developed and maintained software packages (e.g., SAMTools, BCFTools [25], [26]) are problem-specific and therefore do not support the full scope of functionality required for research and diagnostics of rare genetic disorders. The field lacks an open affordable platform providing both completeness of vision and ability to execute.

*AnFiSA* was developed as a computer-aided software[27] with transparent algorithms to facilitate clinical work. It incorporates a novel framework for intuitive and visual application of inclusion and exclusion criteria to millions of records. The framework supports modifications and tuning of the original criteria and provides detailed automatic feedback of how changes in the criteria affect the resulting selection, paving a path to explainable selection models.

While *AnFiSA* is able to support research-based or exploratory analysis of genomic sequencing data, it is built to be integrated into a clinical-grade diagnostic workflow. Thus, it is ideal for providing clinical users with an environment that can help accelerate their research work and efficiently deliver results to downstream clinical applications. Examples of downstream applications include Laboratory Information Management System and Electronic Medical Records. Currently, the new Fast Healthcare Interoperability Resources (FHIR) interoperability framework [28] is the best way to facilitate downstream integration.

To satisfy these requirements, we have based our solution design on the following principles: a) the volume of genetic data should not impact the responsiveness of the system, b) the user interface (UI) should address both research and clinical tasks; and c) the work results should be reproducible and traceable. These principles are translated into a variety of actionable design items:

- A platform that supports building and testing hypotheses in real-time without using programmable languages;
- Importance of real-time inspections of the results at each step of the data evaluation process;
- A platform that can be used as a computer aid for the clinicians;
- A highly responsive system which can deal with billions of genetic variants while providing a sub-minute response time;
- A two-tiered UI with a “red button” tier dedicated to application of existing guidelines and clinical protocols and a research tier supporting flexible research workflow and real-time processing of a patient’s whole genome;
- A visual tool that represents research hypotheses and workflow steps in a readable and editable way, while keeping track of any changes as schematically illustrated by the diagram on Figure 1.

**Figure 1.**
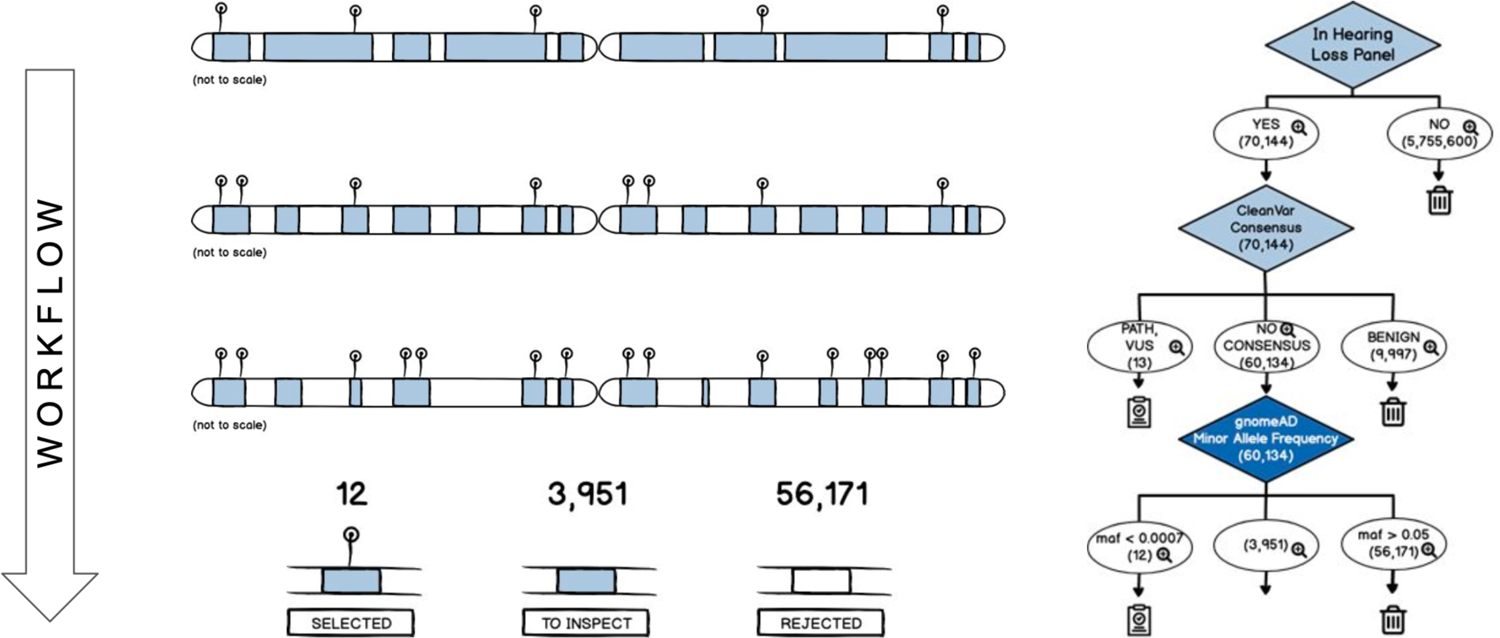
AnFiSA Workflow Ideation

An ideation process to render complex multidimensional data (cubes) and the derived pivot tables (subcubes) in a readable and traceable way. A challenge was to develop a workflow in which each step represents a result of the excluding or including rule operation to funnel the data. This segmentation and an ability to change parameters of each step is best described as the decision tree.

Hence, *AnFiSA* provides a platform simple enough to be used by clinicians while at the same time capable of transforming research workflows into more formal clinical protocols. *AnFiSA* represents these protocols as human readable scripts in a notebook-like environment.

## Design

### Annotation structure

As a warehouse, *AnFiSA* stores annotations of all possible mutations in the human genome that are used to prioritize candidate pathogenic variants in an affected proband. Some of the annotations reflect technical information such as provenance and confidence information about the specific call (call annotations). Provenance data specifies how the call has been made, what tools and data sources have been used and the specific versions of tools or databases. Confidence annotations consist of a set of various call quality characteristics.

Other annotations summarize genetic and biological evidence relevant to the potential effect of mutations on molecular function and phenotype (biological annotations). These annotations combine multiple inputs and consist of genomic, protein, and disease-specific information gathered from different public and proprietary sources. The annotations exist at different genomic scales ranging from the most granular variant-specific annotations to progressively less granular annotations at the level of functional units such as protein domains or regulatory elements, gene level annotations, or annotations of larger genomic segments. Examples of variant-specific annotations include position in a gene transcript, positional sequence conservation in non-human species, and allele frequency of the variant in various human populations.

Beyond granularity of scale, another organizational principle is provided by the knowledge domains contributing the annotations. Human and animal genetics characterize phenotypic effects of sequence variants, various experimental and computational techniques describe the effect on molecular function, and the distribution of variants in human populations is within the purview of population genetics. Annotations are provided by a variety of methods including statistical genetics inference, bioinformatics predictions, and *in vivo* and *in vitro* experiments.

### Annotation sources

*AnFiSA* relies on dbNSFP [29], gnomAD[30], ENSEMBLE Variant Effect Predictor (VEP)[31] and NCBI resources (e.g., ClinVar [32], MedGen and PubMed), OMIM[33], SpliceAI[34] and HGMD as main annotation sources. Collectively, these sources provide information on phenotypic effects of genes and individual variants, allele frequencies, functional effect predictors (e.g., SIFT[35], PolyPhen [36]), conservation scores (e.g. PhastCons[37] and GERP[38]). For ClinVar [32], in addition to the usual data for each variant including clinical significance, stars, review status, conflicts, etc., the user has an option to select a set of trusted submitters, in which case the clinical significance assigned by them will be provided as a separate category. By default, *AnFiSA* includes Laboratory of Molecular Medicine (LMM), GeneDx and Invitae as trusted sources by our team, though designation as trusted can be customized.

The detailed list of annotation sources is provided in the supplementary note S1.

### Phenotypic and inheritance information

The current version of AnFiSA does not include a structured phenotypic information about each case. However, a user can input limited phenotypic data most relevant for the genomic analysis. This includes suspected mode of inheritance and a list of affected and unaffected individuals in the pedigree analysis. The user has the ability to define and interactively explore different inheritance modes by selecting arbitrary groups of affected and unaffected individuals. This feature is important because in many undiagnosed cases, some relatives do not display a clear affected or unaffected phenotype. For the analysis of patient cohorts, AnFiSA supports categorical phenotypic labels; case and control labels are the simplest example.

### Key technology decisions

The main foundation of our development strategy was the realization that genetic data are massive, which makes it impossible to analyze individual records. However, the data are static, which means that they do not change in real time and require analytical rather than transactional processing. The volume of the data naturally leads to the idea of visualization with Pivot Tables.

Pivot Tables show summary statistics and allow rearranging (or pivoting) to spot patterns and build more advanced data models as illustrated in the Supplementary Note S6. They use aggregated values of the properties of the records and present them as various projections of the dataset onto subspaces (usually two-dimensional planes) defined by one or two of the axes (each axis representing a property). In *AnFiSA,* this idea is applied to the process of manual filtration of variants to identify candidate causative mutations when a user builds a filter by adding conditions one-by-one. We believe that, when building and testing clinical hypotheses, the user should have at every step an intuitive understanding of each intermediate subset - the result of application of the already-added conditions. As it is impossible to review individual records in a subset containing many thousands or millions of records, we instead display a pivot table, a widget visualizing the distribution of the properties of the records. Using pivot tables in *AnFiSA* is illustrated in Supplementary note S6. *AnFiSA* does not make decisions for users - rather, it provides them with self-guided tools to navigate in the labyrinth of heterogeneous information.

Genomic data are inherently multidimensional, i.e., every record is described by hundreds of properties coming from a variety of interconnected sources, with each property often having multiple and even inconsistent values often supported by contradicting evidence.

Multidimensional massive data sources are often organized as data warehouses and are processed by application of Online Analytical Processing (OLAP). OLAP generalizes the idea of Pivot Tables to present the data as hypercubes, or, simply, cubes accessible via standardized Database Management Systems (DBMS) interfaces[39]. The term OLAP has been first coined by E.F. Codd in 1993 [40] alluding to Online Transaction Processing - a foundation for traditional DBMS. While Online Transaction Processing balances efficient modification of data with fast access, OLAP tools specifically focus on achieving the maximum performance for data querying and information retrieval [41]. Today, OLAP refers to data management tools that support multidimensional analytical queries and distributed databases. The approach is proven with other verticals like financial analysis, sales forecasting etc., but genetic data have been rarely presented as multidimensional cubes, with a few exceptions described in the literature [42], [43].

Another key concept is representation of complex inclusion and exclusion criteria as a decision tree. This idea has been explored in the past, for example in PASTEL - a platform for assisted clinical trial patient recruitment [44]. Data scientists customarily use computational notebooks to represent their algorithms [6]. Decision trees fit naturally in the notebook interface, where a no-code visual representation of an algorithm is combined with a readable and self-documented script. At each branching point of a decision tree used by *AnFiSA*, variants are split between three buckets: those that are excluded from further review, those that are unconditionally added to the resulting dataset and those that are left for continuation of processing by subsequent branching points.

Finally, there is the idea of crowdsourcing for solving the most challenging clinical cases for patients with undiagnosed diseases [45]. *AnFiSA* gives clinicians an option to create a secondary workspace, which includes only very limited and fully de-identified information and thus can be shared with outside researchers.

### Software Components

As presented in Figure 2, there are several logical elements requiring presentation in the UI.

**Figure 2.**
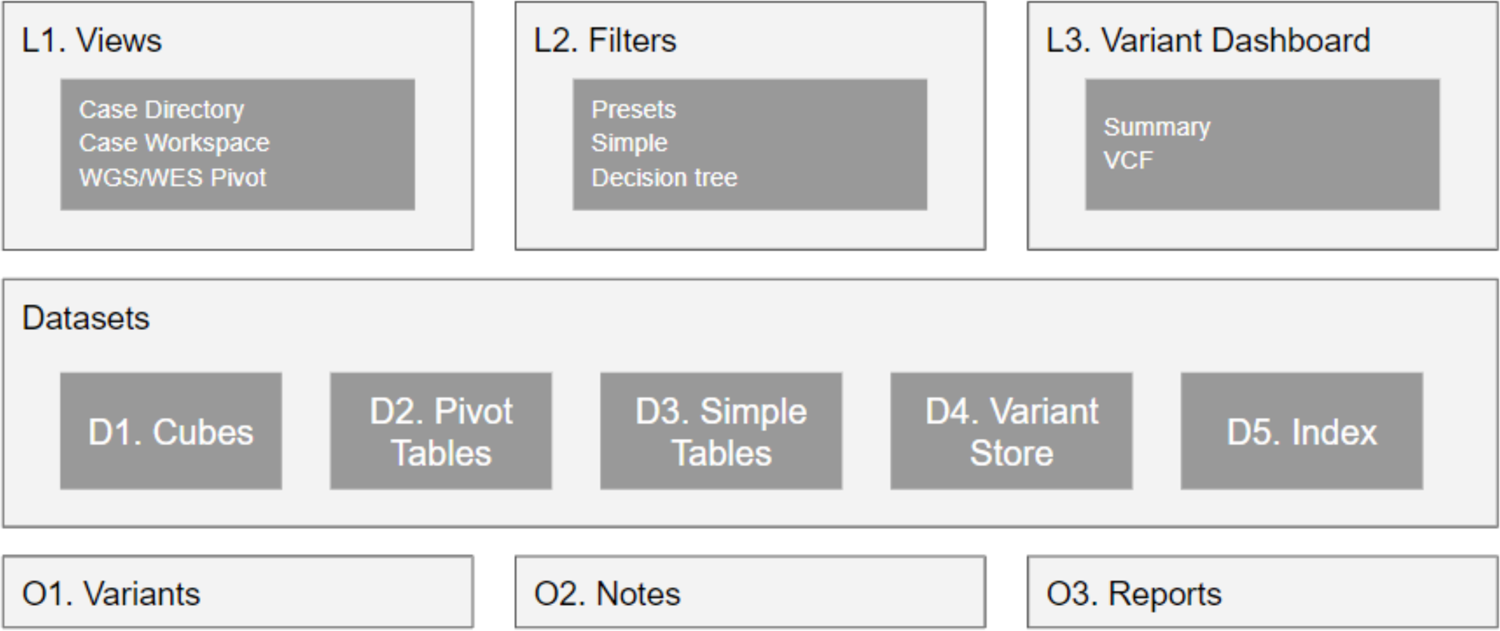
Logical elements of AnFiSA

Several logical elements are designed to address the challenges of curating complex whole genome data sets: views (L1), helping to navigate the data sets, filters (L2) allowing to rapidly minimize the amount of data for manual review, dashboards (L3) facilitating detailed access to individual variants. Datasets comprise a variety of multidimensional structures: datacubes (D1), representing a whole genome or exome, pivot tables built on these cubes (D2), simple tables (D3) storing supplementary materials, variant store (D4) - storing reference data about each variant. Indices (D5) define relations between variants and various kinds of reference data thus facilitating instantaneous navigation in the multidimensional structures. The variants (O1), notes (O2) and reports (O3) are the ultimate work objects presented to the users.

The central component of *AnFiSA* is the backend module developed in Python which interacts with specifically chosen databases as described in the Database Models section below. The end user functionality is exposed through a Representational State Transfer Application Program Interface (REST API). GitHub contains two Restful front-end clients. One is the traditional JavaScript UI used to access new back-end functionality for early adopters. The other is a next generation modern interface developed with Vue.js [46], which supports mature functionality in a more intuitive way.

Along with traditional third-party Extract, Transform, Load (ETL) tools (known as annotation pipelines in bioinformatics), an integrated ETL pipeline was developed as part of the *AnFiSA* project. It is a highly parallel Java program. It creates a feature set for a clinical case by annotating genomes of the patient’s family or a cohort of patients with the data consolidated from various public and commercial databases (provided the end user has a license to specific data).

The modular pipeline is designed to easily plug in third-party annotation tools and functional predictors selected, or even created, by the users (making it easier as most of the community tools are command line Linux programs).

### Data models

There is an ongoing active community discussion of how to model and organize data in medical genomics [28], [47]. Rather than proposing a concrete data model, *AnFiSA* defines basic principles of organization with an ability to accommodate specific data models by provisioning the metadata used for configuration.

Figure 3 illustrates different types of modern database paradigms we have selected for the efficient organization of core genomic data sets, reference data, individual variants, indices, and user generated data.

**Figure 3.**
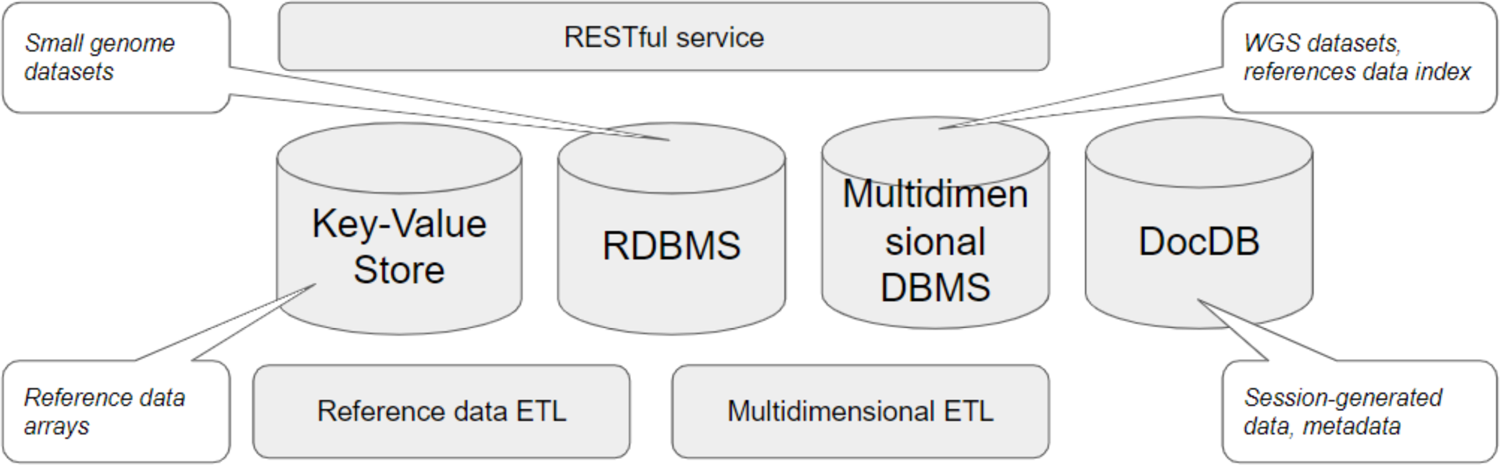
Types of databases to arrange genomic and reference data

A cube in a column-based multidimensional DBMS represents the majority of data from a clinical case. In contrast to a conventional database table, a cube is an appropriate data structure to handle hundreds of millions of records, each characterized by hundreds and even thousands of properties, called dimensions. Multi-value dimensions which we can see in databases such as Apache Druid come handy in genomics where many annotations are ambiguous (e.g. different clinical significance assigned to a variant by different ClinVar submitters). A typical cube for a patient with a family consists of 6 to 7 million records (variants), with each record having from 100 to 2000 non-null dimensions, represented as columns in a multidimensional DBMS. Some cohort-based cases contain dozens of millions of variants. A key-value store contains additional details for each record required for visualization. DocumentDB keeps metadata elements and user generated data originated in UI. In some instances we keep a traditional RDBMS for smaller genomic datasets i.e. gene panels.

### Multi-dimensional modelling with decision trees

The simplest possible model used for variant prioritization is a set of inclusion and exclusion criteria. For example, variants are usually prioritized as potentially pathogenic if their allele frequency in the human population is below a pre-specified threshold and they are coding non-synonymous. A sequential application of inclusion rules is most naturally expressed by a manually constructed decision tree. *AnFiSA* represents this rule-based decision tree as conceptualized in Figure 1.

Specific sets of decision rules may vary depending on the phenotype in question, whether the study is within a research or diagnostic setting, mode of inheritance or simply a preference of the individual researcher/clinician. At the same time, the process is highly repeatable and same sets of rules may be invoked in many similar situations. *AnFiSA* supports a number of pre-defined built-in decision trees. They are included in the distribution and are available in the codebase on GitHub. They can be applied directly or serve as an example for users to build custom decision trees.

Although rule-based decision trees are simple to implement from a conceptual point of view, the sheer amount of data greatly complicates the technical implementation. Inability to host the decision tree in memory requires a solution being a sequence of database queries. To conform to the REST paradigm these queries must be stateless (independent of each other).

*AnFiSA* offered an original solution that is conceptualized in Figure 4, Programming model is described in the Supplementary Note S2, while Supplementary Note S3 provides a syntax reference and Supplementary Note S7 illustrates working with decision trees from a user perspective.

**Figure 4.**
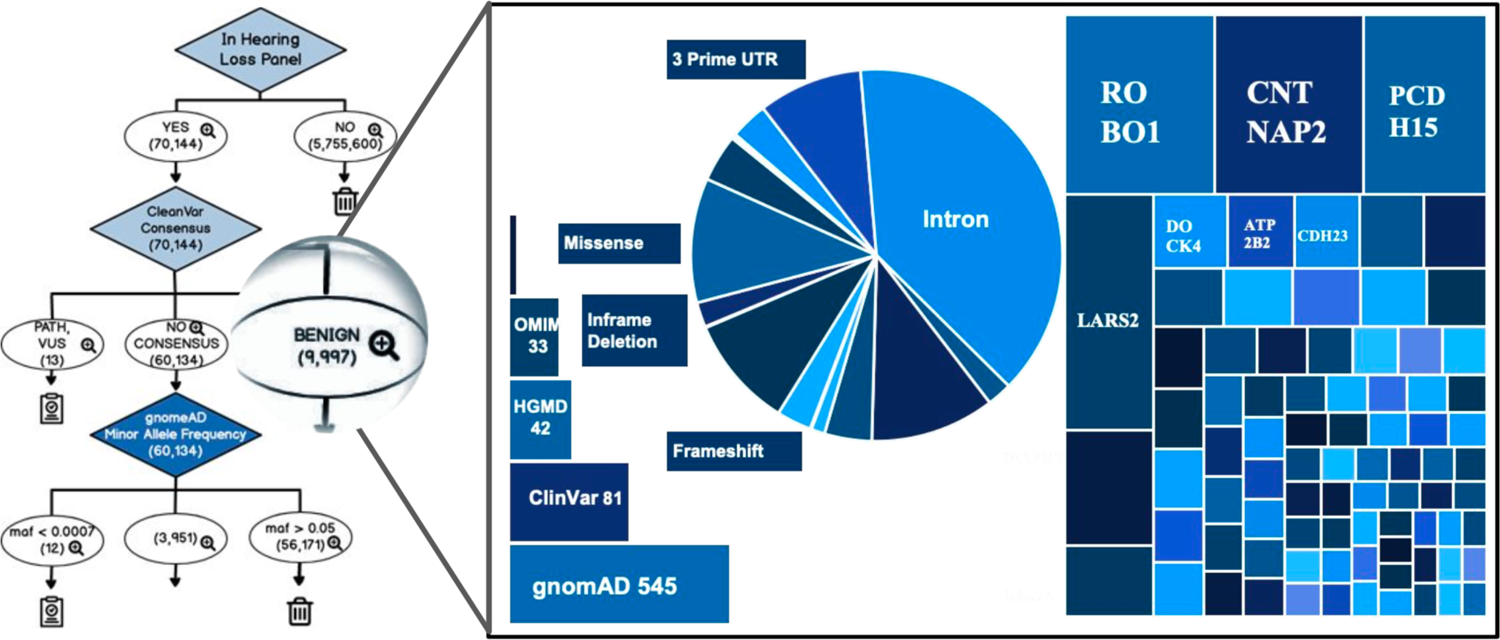
Decision Tree Design Rationale

**Figure 5.**
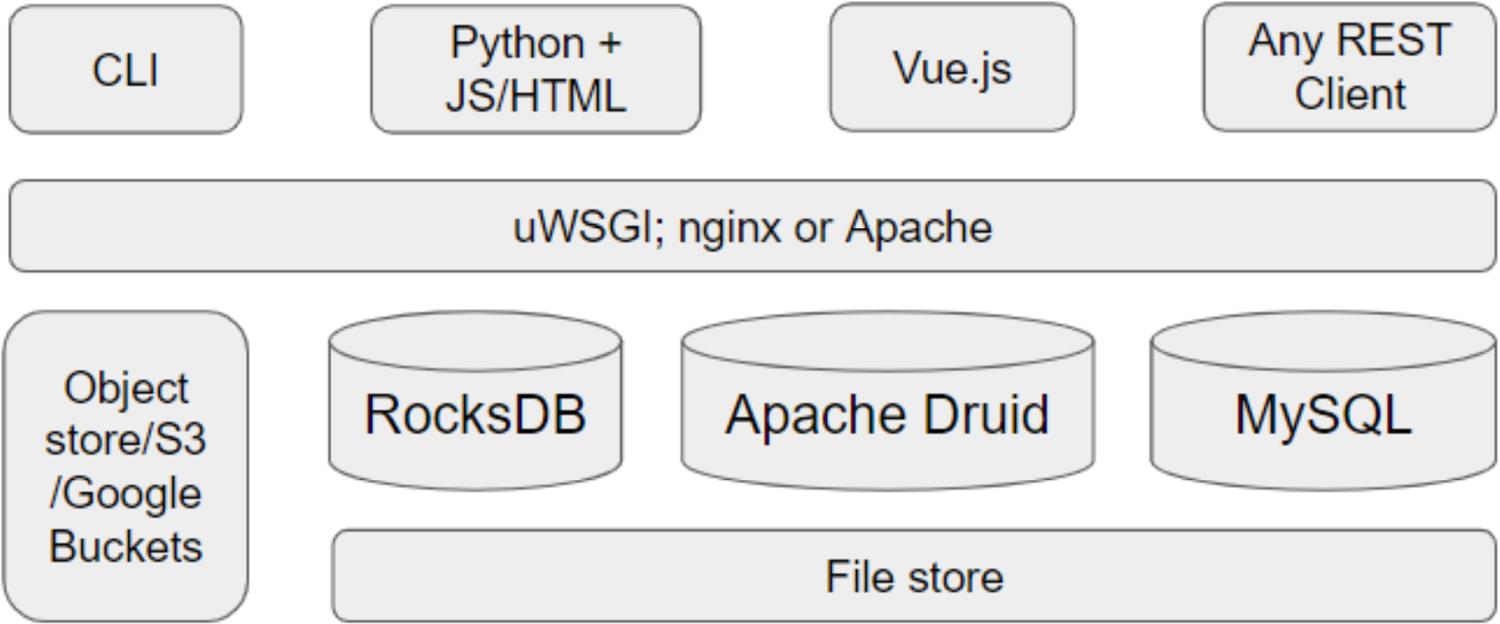
System architecture

A result of application of a decision tree is a set of candidate variants. The starting set consists of all of the variants (in a whole genome of patient family members or a cohort of patients). This set is funneled through a decision tree consisting of a sequence of logical branching points. Each branching point operates on a current set of variants that are split between three buckets: a subset of variants is removed from the set and is either excluded from further consideration (thrown away into a “garbage bin” on the right), unconditionally included in the final selection (on the left) and those that continue their “travel”. Each bucket at each branching point is a sub-cube that can be visualized as a pivot tablee.

### Complex conditions and indexing considerations

One of the advantages of using OLAP is that complex queries can be executed exclusively on indices without accessing actual data records. The selection process is thereby reduced to set operations. This, of course, requires an extensive indexing structure. Apache Druid[48], the current choice for OLAP backend, automatically builds compressed indices for each column (i.e., for each annotation type).

It is important to note that because the set operations are commutative, conditions based on all the above features are also commutative, i.e. they can be applied in any order and yield the same result.

In the world of data warehousing, there are rare tasks that cannot be expressed purely in the terms of set operations because commutativity is lost. Usually, executing these tasks require building specialized indices. In genomics, we encounter non-commutative conditions when we search for compound heterozygous variants, i.e. a combination of two or more heterozygous variants, when one pathogenic variant is inherited from the proband’s mother and another is from the father. We are looking for at least two variants in the same gene.

The result of the application of the compound heterozygous condition and any other condition depends on the order of their application. This can be illustrated by the following example: Suppose that we have selected a set of missense variants in the gene of interest. Now we would like to apply two conditions: that the variants have damaging annotation by PolyPhen and that they are compound heterozygous. If we apply compound heterozygous condition first to identify a pair of variants, and it turns out that one of the variants in the pair is damaging while the other is not, then the subsequent application of “damaging” condition would leave us with one variant. However, if the order of the application of these conditions is reversed, then it will yield no variants.

*AnFiSA* includes a special compound heterozygous variant caller that correctly interacts with other conditions, and this caller will be described in detail in the section “Calling compound heterozygous variants”. Because it uses specialized indices, its mere existence has to be taken into consideration at the time of data warehouse design.

## Implementation

### Testing data

For software development, validation and testing we used data from benchmark variants from the whole genome of the openly-consented “Genome in a Bottle” Ashkenazi trio from the Personal Genome Project [49]. The high-confidence benchmark variants have been provided by NIST version 4.2 [50]. The publicly available demo version of *AnFiSA* includes datasets based on these resources.

### System architecture

As part of our vision focusing on making genomic data accessible, we focus on the data management functionality and encourage the community to develop various UIs for various use cases. We have developed an extending *RESTful API* layer, which gives access to logical objects on our multidimensional model. The persistence layer is primarily provided by a combination of Apache Druid [48] with RocksDB [51]. Our bioinformatics pipeline is illustrated in Figure 6.

**Figure 6.**
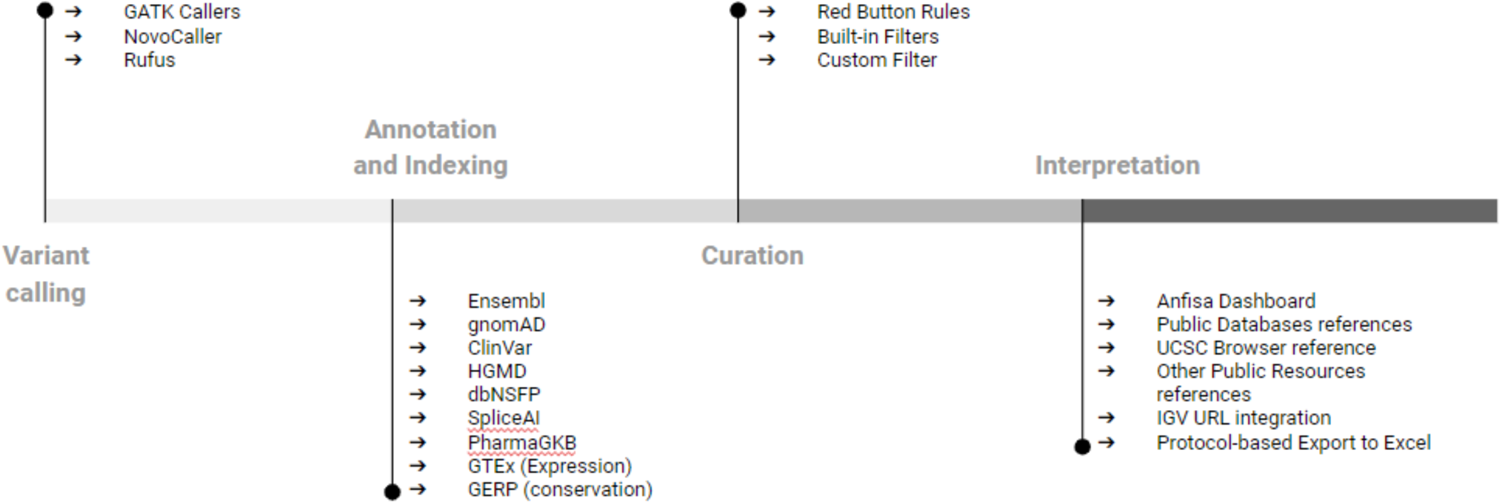
Enhanced pipeline

CLI - command line interface; feature complete experimental UI is based on JS/HTML generated by Python application; Nextgen clinical friendly UI is based on Vue.js; uWSGI is used for serving Python web applications in conjunction with https servers i.e., Nginx and Apache; RocksDB is the key value store for genome variants; clinical cases are stored as a multidimensional model on Apache Druid; MySQL keeps reference data fit for the relational model; *AnFiSA* can be deployed on conventional file systems and various cloud storage types (e.g., S3, Google Buckets).

In comparison to traditional pipelines, our approach suggests additional steps such as indexing to make data accessible for curation with decision trees and other filtering mechanisms. Those additional steps allow a user to visualize only relevant data at each step of designing and adjusting clinical rules. Hence, they are essential to support our key idea of making the variant interpretation process more efficient, transparent and flexible.

### Calling de novo variants

The specificity of calling *de novo* variants by standard GATK callers such as Haplotype caller [52], [53] is unacceptably low to be useful in clinical practice. In many cases, this is not a serious limitation, as *de novo* events are rare and if the analysis is driven by phenotypic features and focused on established disease genes, few *de novo* variants will pass initial filtering. However, *AnFiSA* is designed to be used to evaluate patients with undiagnosed diseases, where *de novo* mutation is often a viable hypothesis. Analyzing such patients requires the use of specialized variant callers. In our practice, we used NovoCaller [54] and Rufus [55]. Calling *de novo* variants is a part of the upstream analysis and hence beyond the topic of this paper. Nevertheless, the curation algorithms we have developed rely on the annotations produced by these callers.

### Calling compound heterozygous variants

Homozygous recessive inheritance occurs when a phenotype is caused by the same variant being present at the same location of both copies of a gene. However, in the case of a loss-of-function phenotype, the recessive inheritance can also occur when two copies of a gene are damaged in different locations. This mode of inheritance is called compound heterozygous.

We have already discussed that a compound heterozygous variant caller is a rare example of non-commutative application of conditions. For this reason, it cannot be run upstream because it takes a set of inclusion and exclusion criteria for the variants as an input. Therefore, it is included in *AnFiSA*.

*Compound Heterozygous Variant Caller* searches for pairs of variants on the same gene that satisfy the following conditions:

1. They are inherited from different parents (*in trans*), and
2. Each variant in the homozygous state can cause loss of function.

When searching for such pairs, it is not sufficient just to look for the consequences’ annotations. Severe consequences for two variants might never manifest together. One example is when a variant belongs to two genes at the same time (for example, when one gene is located within an intron of another). In this case, both variants might be missense, but they are coding for different genes and therefore will not produce the recessive phenotype. More complicated is a case when both variants belong to the same gene but have different annotations for different transcripts. For example, the variant V1 can be in the coding area of transcript T1 and in a non-coding area of transcript T2, while variant V2 is in a coding area of T2 but in a non-coding area of T1. In this case, they will not produce a recessive phenotype either. This consideration is especially important when there is more than one canonical transcript.

Based on these considerations, the primary search for compound heterozygous variants is performed in such a manner that a pair should have matching annotations on the same transcript. This approach is sufficient in the absolute majority of cases. However, in very rare cases of an undiagnosed disease, when there is a strong reason to believe that a variant on a certain gene is causative but such a variant eludes identification, one possible hypothesis might be that a loss of function occurs for a currently unknown transcript. To address this issue, *AnFiSA* allows relaxation of the requirement that matching annotations must belong to the same transcript and look for pairs of variants with matching annotations for the given gene. To facilitate the inclusion of the compound heterozygous criterium in the queries over the whole exome or genome, an index is created on the variant zygosity for each sample.

### Built-in inclusion and exclusion criteria (decision trees)

The following inclusion and exclusion criteria are built-in in *AnFiSA* as Decision Trees:

- An American College of Medical Genetics and Genomics (ACMG) list of 73 genes, in which mutations are known to be pathogenic.
- Rare variants for a trio
- Variants for analysis of undiagnosed diseases
- Variants with damaging predictions
- Possible hearing loss pathogenic variants

The details are provided in the supplementary materials and on GitHub.

### Built-in simple filters in AnFiSA

The following simple filters are built-in:

- Candidate variants for autosomal dominant inheritance
- Candidate variants for homozygous recessive inheritance
- Candidate variants for compound heterozygous recessive inheritance
- Candidate variants for X-linked inheritance
- Variants, possibly affecting splicing
- Variants with *in-silico* damaging predictions

The details are provided in the supplementary materials and on GitHub.

### Deployment

*AnFiSA* has been deployed with on-premises Linux and Mac machines, in the cloud on Linux virtual machines, in a generic Kubernetes cluster and in Red Hat® OpenShift® cluster on IBM Cloud®. It has been tested with OSX 10.14+ and on Ubuntu and CentOS Linux as either a native application or an orchestrated set of docker containers. Docker-compose files are provided for various modes of deployment.

## Results

*AnFiSA* has been used in three diverse and complementary projects at BGM: SEQaBOO, Harvard Undiagnosed Diseases Network [56] Clinical Site, and research cohort analysis of purpura fulminans patients.

### SEQaBOO

SEQaBOO is a clinical research project initiated at Harvard Medical School-affiliated hospitals aimed at integrating high-throughput clinical-grade genomic analysis into routine newborn screening [57]. Using *AnFiSA*, the SEQaBOO clinical team developed a filtration algorithm for the phenotype of congenital deafness. Its application typically identifies 10 to 30 variants from a genome for further review. The *AnFiSA* filtration and annotation functionality was successfully validated by comparison to a hearing-loss analysis process by a separate clinical molecular genetics laboratory and has been incorporated into a CLIA-approved workflow in an established clinical genomics laboratory at BWH. *AnFiSA* has been applied to support a best-practice clinical variant filtration, analysis, and reporting workflow of 40 cases of suspected congenital hearing loss.

### Undiagnosed Diseases Network

BGM uses a phenotype-agnostic approach to analyze rare disease cases refractory to clinical analysis for novel genetic mechanisms of disease [45]. With *AnFiSA*, the search for possible causal variants was performed in two stages. The first stage applies a decision tree to discard variants that are extremely unlikely to be related to the case. The result of this stage is a set of a few hundred variants that are rare, have potential functional significance (e.g., missense) and fit into one of possible inheritance models. In the second step, the clinician drills down into this set, first by applying predefined filters based on the potential inheritance models: autosomal dominant, X-linked and two recessive filters - homozygous recessive and compound heterozygous. If predefined filters yield no obvious result, they can be interactively liberalized or adjusted for alternative inheritance hypotheses (e.g., imprinted genes) to be explored. The compound heterozygous variant caller built into *AnFiSA* allows flexible and interactive combinations of transcript and gene specific annotations into the complex search criteria. Of note, *AnFiSA* version 0.5.16 has now been independently implemented into a cloud-based, end-to-end genomics analysis and interpretation pipeline by the BGM team, which serves as a proof-of-concept for deployment potential. *AnFiSA* has been used by BGM to resolve multiple cases refractory to clinical genomic sequencing analysis, including the case vignette described in **Figure 7**.

**Figure 7.**
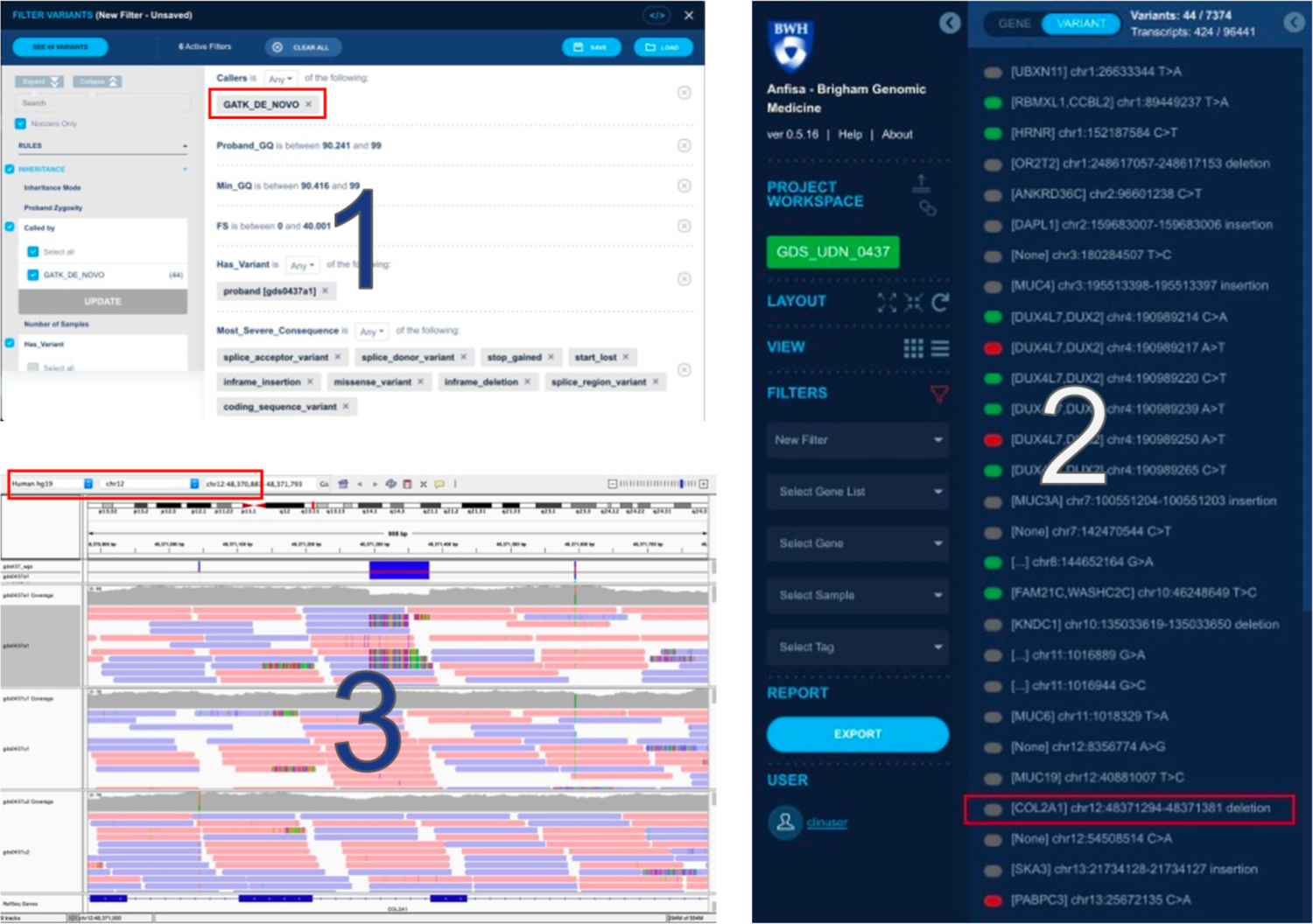
*AnFiSA* Facilitates Identification of Cryptic Genetic Etiology for Stickler Syndrome. 1. Filter Panel: defining criteria to search for candidate variants for autosomal dominant inheritance mode (based on GATK Variant Caller)
2. Candidate Variant Exploration Interface provides a concise list of variants selected by application of the given criteria. Based on phenotypic information, the user selects *de novo* call in *COL2A1*.
3. The Integrated Genomics Viewer, reachable through the embedded link, gives the user the ability to view the BAM file and validate an 87-base intronic deletion that disrupts a canonical splice site.

### Stickler Syndrome Case

Stickler syndrome is a well-defined monogenic disorder typically caused by pathogenic variants in specific collagen genes*: COL2A1, COL11A1, COL11A2, COL9A1, CL9A2, COL9A3.* Clinical synopsis: A female Undiagnosed Diseases Network participant in her 20s with clinical features suggestive of Stickler Syndrome vs. novel inherited connective tissue disease, though the patient had negative clinical-grade gene-panel testing and negative clinical trio WGS (proband, unaffected parents) from reputable commercial and academic clinical molecular genetics laboratories. BGM received raw sequencing files (.fastq) and initiated its WGS pipeline to create a VCF, and then loaded the VCF into the *AnFiSA* annotation and filtration engine. The case analyst filtered and reviewed *de novo* calls from GATK, which include infrequent true positive calls, but are extremely non-specific. However, given the concise list of variants in *AnFiSA*, the user immediately noticed a GATK *de novo* call in *COL2A1*, and using an embedded link for the Integrated Genomics Viewer [58], [59], was able to view the .bam file and validate an 87-base intronic deletion that disrupts a canonical splice site. The variant is absent from the proband’s unaffected parents (see Figure 7). The ability to investigate low confidence variants was critical in this case, because the size of this deletion is beyond the range of the highly specific NovoCaller employed by *AnFiSA* to identify *de novo* point mutations and short insertions and deletions.

The clinical academic laboratory which performed the trio WGS subsequently confirmed the validity of the variant and identified a flaw in their filtration approach that caused the variant to be excluded from manual review. Of note, though this variant was identified on a query of all *de novo* calls from GATK, the analyst was planning to search for variants in the known Stickler syndrome genes using *AnFiSA*’s gene filtration functionality, which would also have identified the *COL2A1* variant. Using *AnFiSA*’s other filters, candidate gene/variant combinations were reviewed for homozygous and compound heterozygous inheritance modes without identification of additional candidates.

### Purpura fulminans

The PF research study uses the cohort analysis capabilities within *AnFiSA*. PF is a dramatic complication of sepsis in which patients experience extreme inflammation, diffuse microvascular thrombosis, and the end organ damage due to uncontrolled activation of the coagulation system. Patients who develop PF tend to be younger than those with more common forms of sepsis and typically have no known acquired or congenital risk factors. These observations strongly suggest the presence of an inherited predisposition to PF[60]. *AnFiSA* was used to investigate the possibility that germline genetic variants contribute to the risk of developing this condition.

A population of 481 samples was divided into three cohorts: PF patients, sepsis patients without PF, and a control cohort. Knowing that both Complement and Coagulation systems play a role in the development of sepsis, *AnFiSA* provided visual comparison of the number of damaging variants in genes involved in both systems for two of the cohorts: sepsis patients who had developed PF and those who did not. Researchers have built filters based on a variant being present in a specific cohort and combination of its frequencies within the cohorts. See Table 1 for additional details on the filtering approach used in this study.

## Discussion

### Software ecosystem for genomic analysis

Development of automation tools for variant the curation process started in late 2000’s.

The first-generation tools have been mostly based on literature search [61]–[65] and had relatively low accuracy. Recently, such tools have moved in the direction of using more structured sources, discovering reference sequences, annotating variants according to ACMG standards, and searching for standard ontology and nomenclature systems of genes and proteins [24], [64]–[67]. Some tools have begun to incorporate machine learning algorithms for the prioritization of variants.

A shared goal for genomic software developers is to create a curation pipeline which utilizes Graphical User Interface operations, and leads to simplified, user friendly NGS data analysis. There is a plethora of tools and platforms for general-purpose data analysis. Top leaders in commercial proprietary data analysis are SAS, Tableau and Microsoft Power BI. On top of this, there are also some well-established free and open-source products like KNIME. Several products, such as Mondrian OLAP Server are open source but intended to be for-profit. On the other hand, many vendors of proprietary software offer free tiers (Oracle Autonomous Database) or academic (SAS, Tableau) or community (InterSystems IRIS) editions of their products that can be used for free.

The diversity of products remains substantial even if discussion is limited to the open-source segment. The most significant platform for open-source projects, ASF, hosts 49 products dedicated to data analysis, not counting products in their incubation stage. Even more products are available outside of ASF. Most of these projects evolve around a community of contributors, where users often offer specific solutions and enhancements to the core product. Many open-source products have multiple forks to create customized versions. Most offer extensive free and commercial support as well as service-level agreements (SLA).

Conversely, the landscape is very different when it comes to genetic and genomic analysis software. A strong indicator for the lack of inclusion of genetic and genomic analysis tools in the software open-source community is that among more than 300 projects hosted by ASF, not one is dedicated to genetics. The majority of clinical Germline Genomic Analysis software is developed by biotechnology companies focused on NGS technology and NGS data analysis, commercial testing labs and some research institutions (e.g., Sherloc [23]). These platforms consist of proprietary software packages. In most cases, it would be a stretch to call these packages as products, as they are not offered to the public, but are used inside genomic testing labs as part of their services. Some organizations claim that their software is open source; however, it is difficult to find these sources and there are certainly no community or external contributors around these organizations.

Curation platforms currently on the market are predominantly developed by hardware vendors or large biotechnology companies providing end-to-end diagnostics services. They offer a wide variety of vendor-specific features, have end-to-end designs, utilize companies’ own proprietary knowledge bases and opaque machine learning and deep learning algorithms for variant prioritization. As discussed regarding functional considerations, these platforms sometimes sacrifice the explainability and traceability of the results[19]–[21]. Genomic clinical laboratories implement score-based variant curation protocols for easier decision making and consequent variant reporting in clinical settings. Multiple curation tools, developed by research institutions, target different steps in curation algorithms, and often operate on genomic variant sets obtained via filtering with the use of third-party tools and programs. Several examples of the currently available curation platforms are included in Supplementary Table ST1.

The landscape of genomic analysis software for academic-based research analysis is quite distinct from the commercial market. Numerous tools have been developed by academic groups. As these tools are usually funded by research grants, they are released as open source and one can easily find their source code on hosting platforms such as GitHub. While technically compliant with the Open-Source Definition, these packages are not fully aligned with the spirit of open-source development. They are not community-supported and do not allow, or at least do not assume, external contributions. Often these tools do not work outside of the specific hardware and environment used by the original developers. In fact, they are not used by anyone except the developers themselves. Many of these tools remain unmaintained once a student or a fellow who has developed them leaves the group. There is no support and no SLA.

There are a few notable exceptions, which are restricted to the tools that are used to solve narrow and very specific tasks, such as genomics format parsers and utilities. BCFTools and SAMTools [25], [26], for example, are well-documented and maintained and come with free support through a number of forums (but still no SLA).

Institutions including the USTAR Center for Genetic Discovery at the University of Utah Health Sciences Center, Stanford University, Washington University in St. Louis and most notably the Broad Institute of Harvard and Massachusetts Institute of Technology, incorporate a conglomerate of the above models by hosting a sizable collection of tools. These collections come closer to representing a comprehensive platform but still fall short because of insufficient documentation and the lack of interoperability guidelines. There is still an absence of, or a very small, outside community of contributors and no real ambition to act as a software vendor. Commercial support and SLAs are usually unavailable.

The organization that comes closest to resembling an open-source community is The Global Alliance for Genomics and Health (GA4GH) - an international, nonprofit alliance formed in 2013 to accelerate the potential of research and medicine to advance human health. According to its statement, the GA4GH community is working to create frameworks and standards to enable the responsible, voluntary, and secure sharing of genomic and health-related data.

GA4GH Cloud Work Stream focuses on API standards. Specifically, Cloud Work Stream develops 4 API standards that allow:

- Sharing of tools/workflows: Tool Registry Service (TRS)
- Execute individual jobs on clouds using a standard API: Task Execution Service (TES)
- Run full CWL/WDL workflows on execution platforms: Workflow Execution Service (WES)
- Read/write data objects across clouds in an agnostic manner: Data Repository Service (DRS).

The claim is that these standards are inspired by large-scale, distributed compute projects which include, for example, PCAWG. Unfortunately, the focus on these standards does not translate into working tools, scoring very low on “ability to execute”.

The implication of the current state of tools for genomic analysis is that if a healthcare organization, such as a teaching or research hospital, wants to establish genetic services based on genomic sequencing, there is no ready-made IT solution that this organization can acquire. The only way is to build a solution internally or to turn to a System Integrator to build a custom solution. Given the rates most System Integrators charge US-based healthcare customers, the cost of IT escalates and becomes comparable with the cost of sequencing equipment and a wet lab. This significantly increases the required investment, and thus slows its adoption.

### Using multidimensional databases for genomics

Genetic analysis belongs to the same class of analytical problems as data warehousing and business intelligence, where relatively static data is processed via application of OLAP. Today, analytical processing is offered by many DBMSs, supporting the Relational OLAP (ROLAP) multidimensional models. Implementations are often based on building in memory cubes and preparing 2-dimensional pivot tables on demand.

Traditional, general purpose DBMS balances fast access to data with its efficient modification and support for granular transactions. Each of these requirements (efficient modification and transaction support) imposes certain constraints on the internal storage organization of the supporting DBMS. For example, all of the data for a logical record should be stored in the same place, preferably within the same physical block on the hard drive. These systems are sometimes called Online Transaction Processing systems.

In contrast, a multidimensional model specifically focuses on achieving maximal performance for complex data querying, data aggregation and information retrieval. It assumes that data modification is performed much more rarely than data queries and that modifications can be done in batch mode, using ETL pipelines. Implementations can handle large amounts of structured and sometimes unstructured data, allowing easy interpretation of big data. Many multidimensional systems use so-called column-based storage, which is especially efficient for data aggregation.

Aggregation is a function on a set of elements. All OLAP engines support simple aggregation functions: element counts, minimums, maximums, sums, arithmetic means, etc. of certain properties of elements in a chosen set. Most OLAP engines also support basic statistical functions like variance. Few support user-defined functions that are supplied as plugins. An example of an engine supporting such plugins is InterSystems DeepSee [43]. The ability to supply user-defined functions for aggregations is an important feature for genomics.

Predominantly, OLAP is used for *ad hoc* queries with a rapid execution time to explore data by viewing specific slices from different viewpoints. In this mode, it is used as a hypothesis generation tool. OLAP is also targeted for decision support and in this capacity, is well-established in many verticals such as financial analysis, sales forecasting, etc. Though genomics data satisfies the condition of being static (more queries than updates and updates in batch mode) and perfectly fits the multidimensional model, to the best of our knowledge OLAP is not used for analysis of massive genetic data sets. Even novel projects that embrace the idea of NoSQL databases overlook the advantages of multidimensional and column-based DBMSs.

Most DBMSs that support the multidimensional model, including Apache Druid [48], also support distributed computing. Utilizing distributed computing can greatly improve response time for complex queries such as cohort building, e.g., when analyzing a large patient population to recruit for clinical trials, run epidemiological studies, etc.

### Future directions

In many ways, the idea of applying curated decision trees for selection of candidate genetic variants was inspired by past experiences of the authors related to development of tools for patient recruitment for clinical trials, as well as for information extraction from free-text medical records. In *AnFiSA* we built an extensive software framework for support of such decision trees. One potential direction is to apply this framework back for patient recruitment and text summarization. Inclusion and exclusion criteria used for clinical trials fit naturally to decision tree model. Many information extraction techniques are based on the presence of certain concepts in either affirmative or negative context, or absence thereof, are expressible as a set of inclusion and exclusion criteria and hence also fit the same decision tree model.

Currently, *AnFiSA* relies on Apache Druid[48] for backend OLAP support. There is an evolution in the data warehousing space, with more and more database vendors having multi- dimensional queries and views as standard features of their DBMS. In the future, we would like to give users more options with enterprise-grade open-source and proprietary products. Among candidates under exploration are Amazon Redshift, Oracle Autonomous Data Warehouse and IBM Cloud Pak® for Data.

Another direction in the development of *AnFiSA* is providing a more streamlined ability for the user to build explainable predictive models other than the decision trees, including supervised and unsupervised machine learning and deep learning models. Recently, multiple research groups attempted advancing Explainable Artificial Intelligence (XAI)[68], [69] in two directions. The first is explaining black box models by either computing the sensitivity of the prediction with respect to changes in the input (e.g., [70] or decomposition of the decision in terms of the input variables (e.g., [71]). The second is building white-box, interpretable machine learning models, that come with their own explanations [72]. *AnFiSA* data model is designed to suite both directions. Despite significant challenges [73], the sheer volume of recent publications and workshops in XAI give a hope that explainable deep learning models can be in the future used in medical diagnostics in general, and genetic diagnostics in particular. Incorporating these techniques into *AnFiSA* is certainly our priority.

As referenced in the Application section, *AnFiSA* can be used in patients’ cohort analysis. As more complex cohort analysis requirements mature, we are going to explore the value of a distributed computation to balance the query load between nodes. This is an out-of-the-box capability of many OLAP implementations in modern DBMS such as Apache Druid and Apache Hadoop.

Finally, another important development direction is computer-aided report generation. Currently a user can provide an Excel template to export the selected variants into an Excel Workbook. We anticipate adding the ability to provide Microsoft Word templates to generate a CLIA certified report.

## Supplementary materials

See SupplementaryMaterials.docx

## Data Availability

Source code is available on GitHub.

https://github.com/ForomePlatform/AnFiSA#public-demo

https://github.com/ForomePlatform

