## Supplemental_material for "AnFiSA: An open-source computational platform for the analysis of sequencing data for rare genetic disease"

### Supplementary Materials

Supplementary Note S1: Annotation Sources 2
Supplementary Note S2. Decision Tree Programming Object Model 4
Supplementary Note S3. Syntax for Inclusion and Exclusion Criteria 5
Decision Tree Logic 5
Syntax Principles 5
Decision Tree Syntax Reference 6
Decision Tree system support 7
Supplementary Note S4. Variant Classification 9
Supplementary Note S5. Anfisa Home Screen 10
Supplementary Note S6. Exploration of Pivot Tables 11
Supplementary Note S7. Working with Decision Trees 13
Supplementary Note S8. Variant curation within a derived dataset 15
Phenotype-based analysis (looking for potential hearing loss variants) 15
Genetics first/Phenotype agnostic analysis 16
Supplementary Note S9. Reporting chosen variants 18
Manual review and tagging of the variants 18
Export options 18
Supplementary Note 10. GitHub Repositories Structure 19
Code Sample. Built-in Curation Rules 20
Inclusion and Exclusion Criteria 20
Simple Filters 25
Supplementary Table ST1. Examples of Available Variant Curation Platforms 27
Supplementary Table ST2. Color and Shape codes used for variants visualization 28
Supplementary Table ST3. Ratios of Damaging to Benign Variants for Sepsis Patients with and without PF 29

### Supplementary Note S1: Annotation Sources

#### Assemblies

**GRCh37 (hg19)**

- Project URL https://www.ncbi.nlm.nih.gov/assembly/GCF_000001405.13/
- File URL: http://ftp.ncbi.nlm.nih.gov/genomes/refseq/vertebrate_mammalian/Homo_sapiens/all_assembly_versions/GCF_000001405.25_GRCh37.p13/GCF_000001405.25_GRCh37.p13_genomic.fna.gz

**GRCh38**

- Version: patch 13
- Project URL: https://www.ncbi.nlm.nih.gov/assembly/GCF_000001405.39
- File URL: ftp://ftp.ncbi.nlm.nih.gov/genomes/refseq/vertebrate_mammalian/Homo_sapiens/all_assembly_versions/GCF_000001405.39_GRCh38.p13

#### Gencode GTF

- Project URL: https://www.gencodegenes.org/pages/data_format.html
- Downloads: https://useast.ensembl.org/info/data/ftp/index.html
- Direct Download URL: ftp://ftp.ensembl.org/pub/release-99/gtf/homo_sapiens/Homo_sapiens.GRCh38.99.chr.gtf.gz

#### Genome Aggregation Database (gnomAD)

- URL: https://gnomad.broadinstitute.org/
- Download URL: https://gnomad.broadinstitute.org/downloads
- Version: 2.1.1

#### ClinVar

- Project URL: https://www.ncbi.nlm.nih.gov/clinvar/
- CSV File URL:https://ftp.ncbi.nlm.nih.gov/pub/clinvar/tab_delimited/variant_summary.txt.gz (contains data)
- XML File URL (contains data and metadata): https://ftp.ncbi.nlm.nih.gov/pub/clinvar/xml/

#### SpliceAI

- GitHub URL:
- Download URL (requires free registration): https://basespace.illumina.com/analyses/194103939/files/236418325?projectId=66029966
- Version: v1pre3
- Files:
    - spliceai_scores.masked.snv.hg38.vcf.gz
    - spliceai_scores.masked.indel.hg38.vcf.gz
- https://github.com/Illumina/SpliceAI

#### dbNSFP

- Project URL: https://sites.google.com/site/jpopgen/dbNSFP
- File URL:
  - ftp://dbnsfp:dbnsfp@dbnsfp.softgenetics.com/dbNSFP4.0a.zip
  - https://drive.google.com/file/d/1BNLEdIc4CjCeOa7V7Z8n8P8RHqUaF5GZ/view?usp=sharing

#### GTEx

- Project URL: https://www.gtexportal.org/home/
- Download URL: https://storage.googleapis.com/gtex_analysis_v8/rna_seq_data/GTEx_Analysis_2017-06-05_v8_RNASeQCv1.1.9_gene_median_tpm.gct.gz

#### PharmGKB (Pharmacogenomics)

- Project URL: https://www.pharmgkb.org/
- Download URL: https://www.pharmgkb.org/downloads
- File: Variant Annotations Help File (annotations.zip)

#### GERP Scores

- Project URL: http://mendel.stanford.edu/SidowLab/downloads/gerp/
- Download URL: http://mendel.stanford.edu/SidowLab/downloads/gerp/hg19.GERP_scores.tar.gz

#### Supplementary Note S2. Decision Tree Programming Object Model

The internal object model for decision trees includes the following classes of objects: atomic condition (i.e., atoms), instructions and named states. There are two types of instructions: those that calculate values, and those that select into which bucket the result should be deposited. Values are most often calculated using complex conditions, i.e., logical combinations of atoms, but in rare cases can be calculated by calling a plugin function. At execution time, each instruction is related to a subset of data. In case of calculation instruction, it is a subset to which condition or function is applied. In the case of bucket selection instruction, it is a subset of records being put into the selected bucket. These subsets are in fact sub-cubes and can be visualized as pivot tables as conceptualized by the diagram in Figure 4. Named states are a more advanced entity and can be used if a calculation needs to be performed on a dataset different from the current one, just before a bucket selection instruction. They are used by the compound heterozygous variant caller.

A decision tree can be projected to the user either as a script in a dialect of Python or as a combination of widgets used by the UI. Both projections are exposed via REST API. Due to complexity of the objects representing decision trees, the backend provides abundant metadata about them, like a markup for syntax highlighting of a script. Metadata is also provided through the REST API. The built-in UI client allows users to either type or edit the script for a decision tree or build/change the tree interactively. A rule represented as a decision tree is transparent and replicable, and its representation in a scripting language can be easily included in any protocol.

A typical decision tree consists of about 15 nodes, with each node representing a complex condition on the functional, clinical, sequencing quality, call quality and/or population genetics data. In addition to allowing a user to view and edit a condition at each node, the UI gives the researcher or clinician a runtime capability to easily examine the variants at that node and review which variants have been put into which bucket. Therefore, a user who is developing a new rule can easily see any mistakes or inefficiencies and improve upon or optimize that rule. Application of a rule to an annotated and indexed whole genome (about 6 million variants) typically takes about one minute and produces a set of between a dozen and a few hundred variants that are presented to a clinical geneticist for subsequent manual review.

### Supplementary Note S3. Syntax for Inclusion and Exclusion Criteria

#### Decision Tree Logic

A Decision Tree consists of a sequence of branching points. A result of application of any Decision Tree is a set that we will call "final selection". The process starts with a set consisting of all the variants (in a whole genome of patient family members or a cohort of patients). This set "travels" through a tree trunk. At each branching point a subset of variants is removed from the set and is either excluded from further consideration (thrown away) or unconditionally included in the final selection. Variants that have been neither excluded nor included, continue their "travel".

- Initially we have the whole set of items (variants) as working selection.
- At each branching point:
  - If-instruction selects some subset of working selection;
  - Return-instruction determines whether the selected subset should be included in the "final selection" (return True) or excluded (return False):

**if** *condition*:
    **return** *bool decision*

- after If-instruction the selection set is (probably) reduced, and next instruction is applied to this reduced set; next instruction is one more If-instruction, or…
- final instruction in code is always Return-instruction that determines what should be done to the rest of working selection: to include it in the "final selection" (True) or to exclude it (False):

**return** *bool decision*

There is only one other type of available instruction, Label-instruction:

**label** (*string*)

This instruction can be inserted to decision tree code before any If-instruction. So the user has a possibility to mark the state of working selection by label mark. This mark can be used in complex procedures (see functions reference: Filtering functions, functions Compound_Heterozygous() and Compound_Request()).

#### Syntax Principles

There are three levels of details in description of Decision Tree Python dialect:

- **necessary level**: the dialect deals with very restricted subset of Python, so only a small subset of Python constructs is allowed; below is complete description of this subset

- **good practice level**: some constructs discussed below are recommended as "good practice"; similar constructs that are not considered good practices could be refactored to their "good practice" analogues in the process of interactive changes of a decision tree

- **simplification level**: since the dialect of Python is very "thin", for purposes of easy typing and reading it supports the following "simplifications":
  - **string constants** can be typed without quote symbols `""` or `'` if they are correct Python identifiers or constants `True, False, None`
  - **lists vs. sets**: in case when code refers list objects with `[]` parentheses, it is good practice to use set notation with `{}`; indeed, in most cases, order of elements in a "list" is irrelevant, while `{}` are more readable

#### Decision Tree Syntax Reference

##### Top level constructs

There are three top level constructions available in the dialect:

**if** *condition* :
**return** *bool decision*

**return** *bool decision*

**label** (*string*)

The following rules must be hold:

- All instructions (excluding Return-sub-instruction of If-instruction) must start at the first character of a line, no indentation

- A top-level Return-instruction must be the last non-empty line of code
- Label-instruction can be used before any If-instruction
- Empty lines between top-level constructions are allowed
- Comments are acceptable only as a full line, not as a part of a line with code; comments should start with `#` character, possibly after spaces (note also that comments are not acceptable after the last instruction)

- It is a good practice to place comment lines only before top-level instructions
- *condition* in If-instruction might be quite long, so one might need multiple lines; It is good practice to use parentheses to group these lines, instead of `\` characters.

##### Conditions

**Combined conditions**

Operators `and`, `or` and `not` and parentheses `()` are fully supported for building complex conditions from atomic ones.

Atomic condition uses an identifier of corresponding filtering property once per atomic condition. (See also Condition descriptor for understanding atomic operations.)

**Atomic numeric condition**

Has form of usual Python comparison operation with operators `<`, `<=`, `==`, `>=`, `>`. Double form is acceptable, for example:

*min value < property_id <= max_value*

Best practice: use only operators `<`, `<=`, `==`; in case of operator `==` place property identifier on the left.

**Atomic enumerated condition**

Has different form in dependency of join mode of condition:

**OR**:
*property_id* **in** { *set/list of value strings* }

**AND**:
*property_id* **in all** ({ *set/list of value strings* })

**NOT**:
*property_id* **not in** { *set/list of value strings* }

Notes:

- notation above uses `{}` set parentheses; though it is recommended as a good practice,

  list parentheses `[]` are also supported
- operator **in** is supported for all enumerated properties, including status (single-value properties) and multiset (multi -value properties). Semantic of status properties is simple and intuitive.

  In case of multiset properties, this notation is more sophisticated: the condition is positive when intersection of two sets is nonempty, i.e. at least one value of the property matches at least one value in the given set; it can be "explained" by a way that object representing filtering property redefines operator **in** from the left
- in case of **AND** join mode interpretation of **all()** pseudo-function is

  even more sophisticated: it can be "explained" if result of **all()** redefines" **in** operation in a very specific way from the right.
- in terms of Decision Tree there is no strong need for **NOT** join mode,

  because operator `not` is supported outside atomic conditions

**Atomic function conditions**

Function conditions have similar form to enumerated conditions with a change of *property id* to *function_name* (*parameters*)

Syntax for parameters is Python standard. Since all values of the parameters must be JSON objects (however, with a change of JS constants true/false/null to Python counterparts True/False/None), there should be no problems in setting parameters up. ("Simplifications" are also acceptable for parameters).

See Filtering functions for reference of available functions and their parameters.

#### Decision Tree system support

The following objects are explicated from the code of decision tree:

- **Points** correspond to instruction in code; each If- or Return- instruction corresponds to a point with state of selection set: either working one or pre-final. The user needs to know how many items (variants) are in these sets, and moreover, has a possibility to study distribution of values for filtering properties of items in these sets.
- **Atomic conditions** are "atomic" fragments of condition in If instructions. There can be many atomic conditions in one If instruction. It is important functionality of the system to locate them and provide their modifications.
- **State labels** can be defined in code by Label instructions. They are used with complex functions. This functionality requires a high level of qualification and attention of the user; however they might be very important in practice.

A decision tree can be modified in either of two ways:

- manual typing and modifications of decision tree code
- interactive actions modifying various details of decision tree, see Decision Tree modifying actions for reference.

Interactive regime allows to make any meaningful transformation of a decision tree, so there is no strong need to use manual regime at all. Manual regime requires is helpful for complex manipulations with boolean logic of conditions and, of course for copy/paste operations.

#### Supplementary Note S4. Variant Classification

Anfisa provides users with the ability to semi-automate variant classification with a transparent decision model. Clinical classification of the sequence variants is calculated by a combination of intermediate parameters. The parameters are comprised of information programmatically assessed from various public databases, which include variant consequences, clinical significance allocated for the variant by HGMD and ClinVar databases and use of the genetic *in-silico* prediction tools, such as PolyPhen2, SIFT, FATHMM, Mutation Assessor and Mutation Taster.

The variant's consequence is divided into two groups identified either as a putative loss-of-function variant or as unknown functional impact. By default, this grouping is the first step in a variant's classification in Anfisa. The second step is filtering against the tags assigned to the variant in HGMD and ClinVar databases. The four values (consensus benign, consensus pathogenic, uncertain predictions and absent predictions) could be assigned to the variants. Consensus benign or pathogenic values are assigned to the variants for which HGMD and ClinVar tags are either concordant in both databases or have been tagged as consensus benign or pathogenic at least in one of the databases. An uncertain prediction value is assigned for the variants with discordant HGMD and ClinVar tags. For the variants with no consensus benign or pathogenic values, or variants that are absent from both databases, *in-silico* prediction tools are applied as a third step in the variant's classification. Supplementary Table ST2 in the supplementary materials illustrates the variant classification algorithm.

Corresponding to the variant classification, visual labels are assigned as crosses for variants leading to loss-of-function and as circles for the variants which do not implicitly disrupt function of the protein coding genes. The pathogenicity of the variants is coded by color. Benign variants are colored green, variants of uncertain significance are colored yellow, and pathogenic variants are colored red. However, in case of *loss-of-function*, benign variants are colored yellow despite being classified as not damaging due to nature of the variant consequence. Sequence variants which are not listed in HGMD and ClinVar and do not have *in-silico* predictions are displayed as gray circles.

#### Supplementary Note S5. Landing page with datasets

Analytical processing, annotation and loading of a whole genome into Anfisa Backend is a back-office operation. Therefore, in practice, user experience starts when a clinician is presented with a whole genome dataset in *Anfisa* UI. The core of the dataset is a datacube, i.e., a multidimensional array of values accessible via standardized DBMS interfaces [54] representing the genetic variants. In addition, each dataset includes various supporting data and documentation (Figure SF1). All datasets provide information about the tools and their versions that have been used to process the data with the date when the tools have been executed. Additional documentation can include clinical notes, quality control and other reports. *Anfisa* handles the following formats for supporting documents: plain text (.txt), html and various image formats (.jpeg, .tiff, .png, etc.). Which specific documents are shown depends on the upstream pipeline and on how *Anfisa* is configured. For example, BGM and *SEQ*uencing a Baby for an Optimal Outcome (SEQaBOO) implementations of *Anfisa* provided the following documentation: quality control (QC report, coverage histograms, ancestry principal component analysis plots, callability reports), reports from copy number variation analysis, and reports generated by a virus detection pipeline.

***Figure SF1. Landing page with genome datasets***

(1) Dataset name; (2) case documentation; (3) quick access to inclusion/exclusion panel

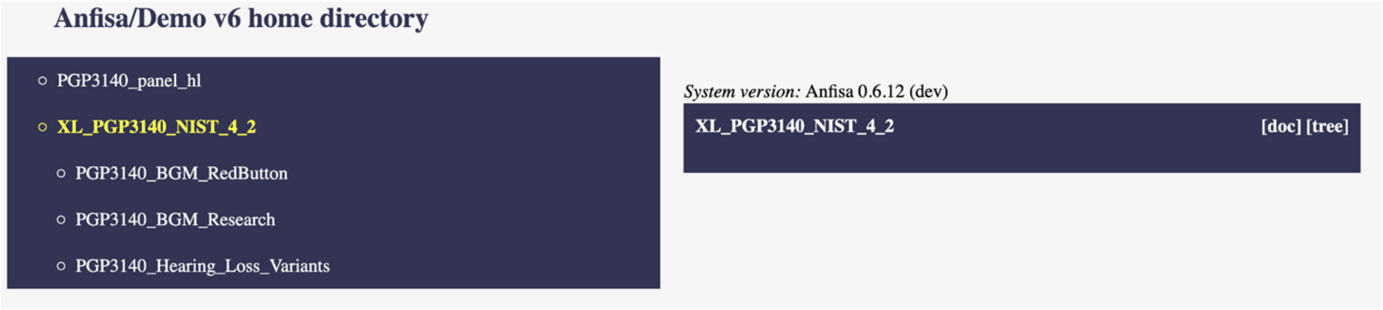

#### Supplementary Note S6. Exploration of Pivot Tables

It is often convenient to explore the number and basic properties of variants selected according to various combinations of filters without analyzing these variants individually. For example, a user may wish to quickly find out how many coding variants are present in the proband's genome or how many of these variants have population frequencies below a given cutoff. Anfisa presents a summary of pivot tables that includes the number of variants in a subset, the ranges for all variant property values, and the distributions of variants in the value range.

We illustrate this process in the study of Purpura fulminans (PF) patients [53]. These steps are illustrated in Figure SF2.

##### *Figure SF2. Pivot Exploration*

Exploration Interface for the PF dataset. (1) The jointly-called whole exome data set contains 1,577,451 variants. (2) The user adds a condition on allele frequency from gnomAD, to review only the variants with allele frequency less than 0.05. The number of variants is now reduced to 1,368,827. (3) The user selects variants from the Complement System Gene Panel, reducing the number of variants to 2,785. (4) The user selects only variants that are found in patients from the PF cohort, which brings the user to 1,331 variants, scrolls down to the in-silico predictions section, and records the numbers of damaging and tolerated variants. Then the user changes the cohort to Sepsis (control).

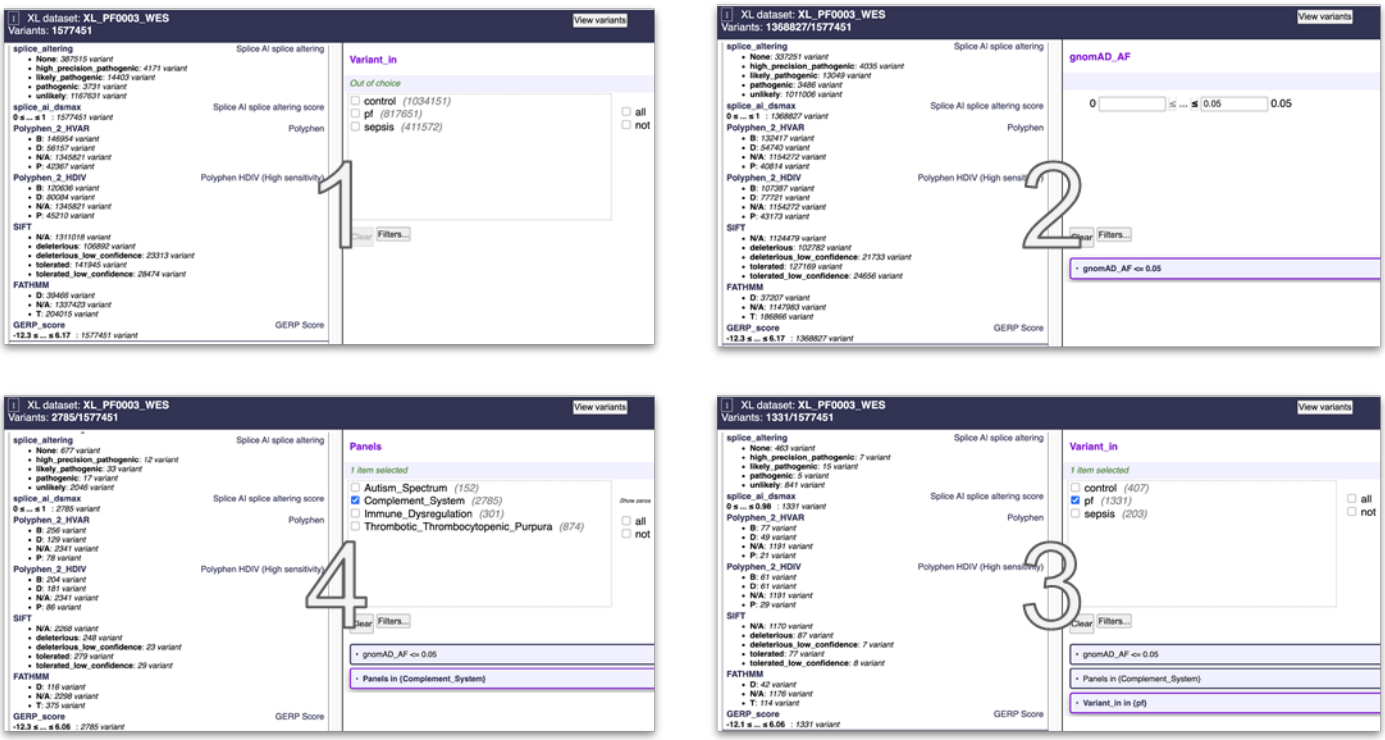


The next step is to change the gene panel to **Coagulation System Gene List** and compare PF and Sepsis cohorts. The exploration results are presented in Supplementary Table ST3.

While the information obtained through the process described above cannot be used as evidence, it can provide a hypothesis that can subsequently be verified through rigorous statistical analysis. It should be noted that at no point in this procedure have we looked at individual variants (which is the next stage of the research process), but instead used pivot tables.

#### Supplementary Note S7. Working with Decision Trees

In *Anfisa*, the recommended first step of variant curation is the application of a high sensitivity filter to select a large set of putative variants, or rather to exclude variants that we are confident are irrelevant. The filter represents an inclusion/exclusion criterion and is usually implemented as a decision tree (Figure SF3). The operation corresponds to a database query and yields a compact subset of variants. The subset can be saved as a derived dataset, like construction of a materialized view in a Relational DBMS. In the current version of *Anfisa*, this is an explicit operation triggered by the user when selecting the "Save as derived dataset" operation from the menu. We anticipate that in future versions, it will become an implicit operation, like automated gear shifting in cars with automatic transmission.

##### *Figure SF3. Inclusion and Exclusion Criteria.*

Inclusion/exclusion criteria is implemented as a decision tree. Each block is a branching point. A condition shown in the center panel of the window is applied to a set of variants considered at the given step. Based on the condition, the variants are either ultimately included in the candidate list (shown with green "+" sign in the left panel) or excluded from further consideration (shown with brown "-" sign in the left panel). The right panel is a pivot table corresponding to a set of variants at a given step: under consideration, included or excluded. Each of these sets can be examined for more details.

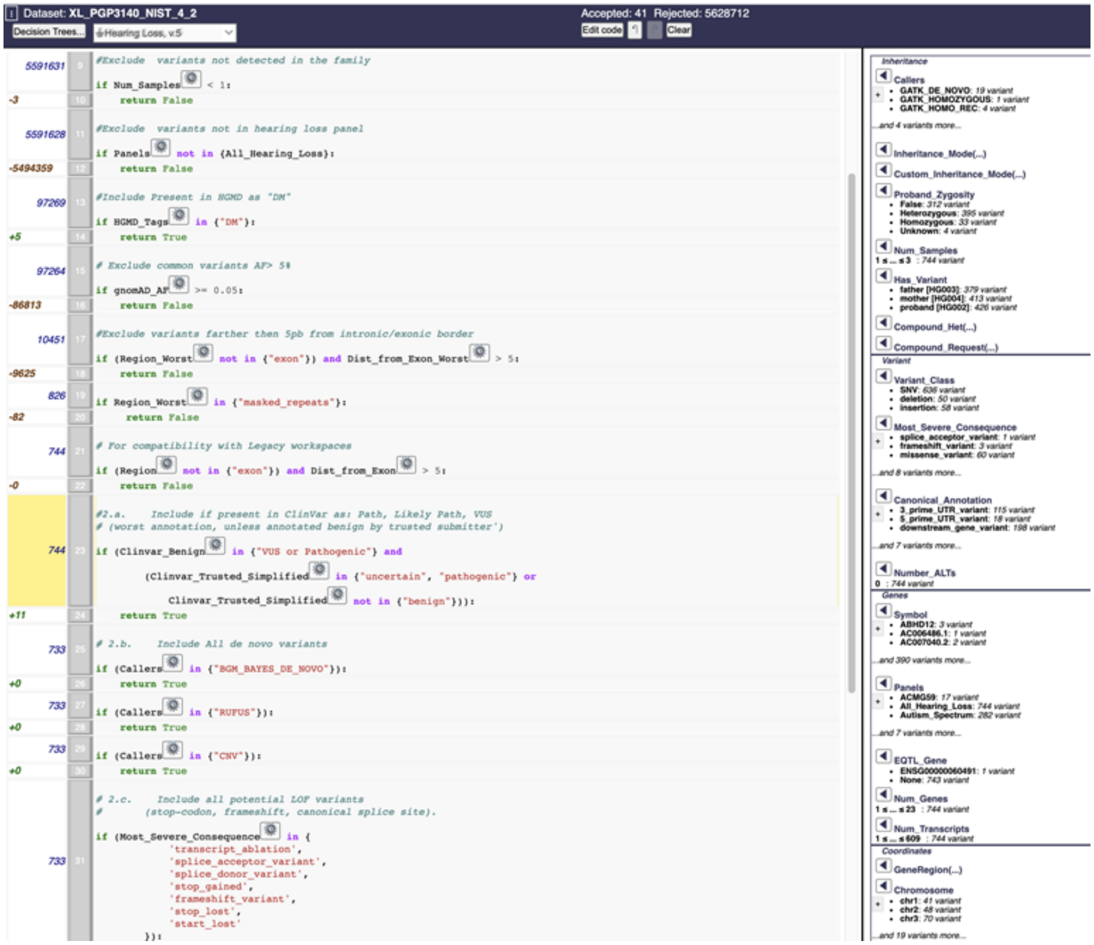

The inclusion/exclusion criteria can either be based on phenotypic information, when potentially damaging variants are selected from a user-defined list of genes, or on the "genetic first" approach [40], which uses a certain inheritance model consistent with the observed phenotypes. The *Anfisa* code base provides examples of decision trees for both approaches. The demo version of *Anfisa* includes benchmark variants from the whole genome of openly-consented "Genome in a Bottle" Ashkenazi trio from the Personal Genome Project [55]. We use the high-confidence benchmarks provided by NIST version 4.2 [56].

From a datacube representing the whole genome, a user can create multiple derived datasets corresponding to different working hypotheses. In the demo version of *Anfisa* we provide three derived workspaces based on the NIST high confidence variants: one for hearing loss candidate variants, one based on the BGM "Red Button" criteria, and one based on the BGM research criteria.

#### Supplementary Note S8. Variant curation within a derived dataset

Each derived dataset provides the same functionality as a datacube with two additional features:

1. Variants can be directly visualized and manually reviewed.
2. More granular inclusion/exclusion criteria based on the selection of individual transcripts.

For example, it is possible to pick variants that are selectively predicted to be damaging only in Ensembl transcripts or in a specified version of RefSeq transcripts. This is an important functionality for searching for compound heterozygous variants.

During data transfer of the selected variants set into a derived dataset, each variant is classified according to ACMG guidelines, and assigned a color-coded label.

##### Phenotype-based analysis (looking for potential hearing loss variants)

A sample workflow for hearing loss can be illustrated using a demo dataset derived from the "Genome in a Bottle" Ashkenazi trio NIST v4.2. Because we are using public data, the proband has normal hearing, so the following workflow is for illustration purposes only. A user can either create a derived dataset by applying the "Hearing Loss v.5" decision tree or by using a prebuilt dataset named PGP3140_Hearing_Loss_Variants.

The dataset contains 41 variants. Inside the dataset, the user first applies the "Hearing Loss Quick Filter", which implements the logic based on the selection of individual transcripts to further reduce the number of variants to 24. The filter itself can be examined in the Decision Tree Panel. Apart from the variants found in the proband, the 24 variants include the carrier variants found only in parents. The user should exclude those variants by selecting "proband" in the "Sample" drop-down menu. That leaves 8 variants. The user then reviews these variants manually.

The first variant is chr4:54727298 A>C in *KIT* (Figure SF4). It is included in the dataset because it has the "DM" tag in HGMD. However, it is a common variant in the Ashkenazi Jewish population to which the proband belongs and is annotated as "Benign" in ClinVar. A clinician would likely exclude this variant from consideration. In *Anfisa*, this is done by tagging a variant as "Likely Benign" and leaving a corresponding note.

***Figure SF4. Looking at KIT chr4:54727298 A>C Variant***

We illustrate working in a derived dataset on the PGP3140_WGS_HLPANEL dataset created for a custom hearing loss panel. The full dataset contains 2529 variants. After application of "SEQaBOO Hearing Loss v.3.5" preset filter (top left lis-box) a user is presented with 21 putative candidate variants. Selecting variants present in proband ("Select Sample" list-box further below) leaves the user with 11 variants.

The user then reviews these variants manually. We will look in details at the variant chr4:55569954 A>C (chr4:54703788 A>C in HG38) in *KIT*. It is included in the dataset because it has the "DM" tag in HGMD. However, it is a common variant in the Ashkenazi Jewish population to which the proband belongs and is annotated as "Benign" in ClinVar. A clinician would likely exclude this variant from consideration. In Anfisa, this is done by tagging a variant as "Likely Benign" and leaving a corresponding note.

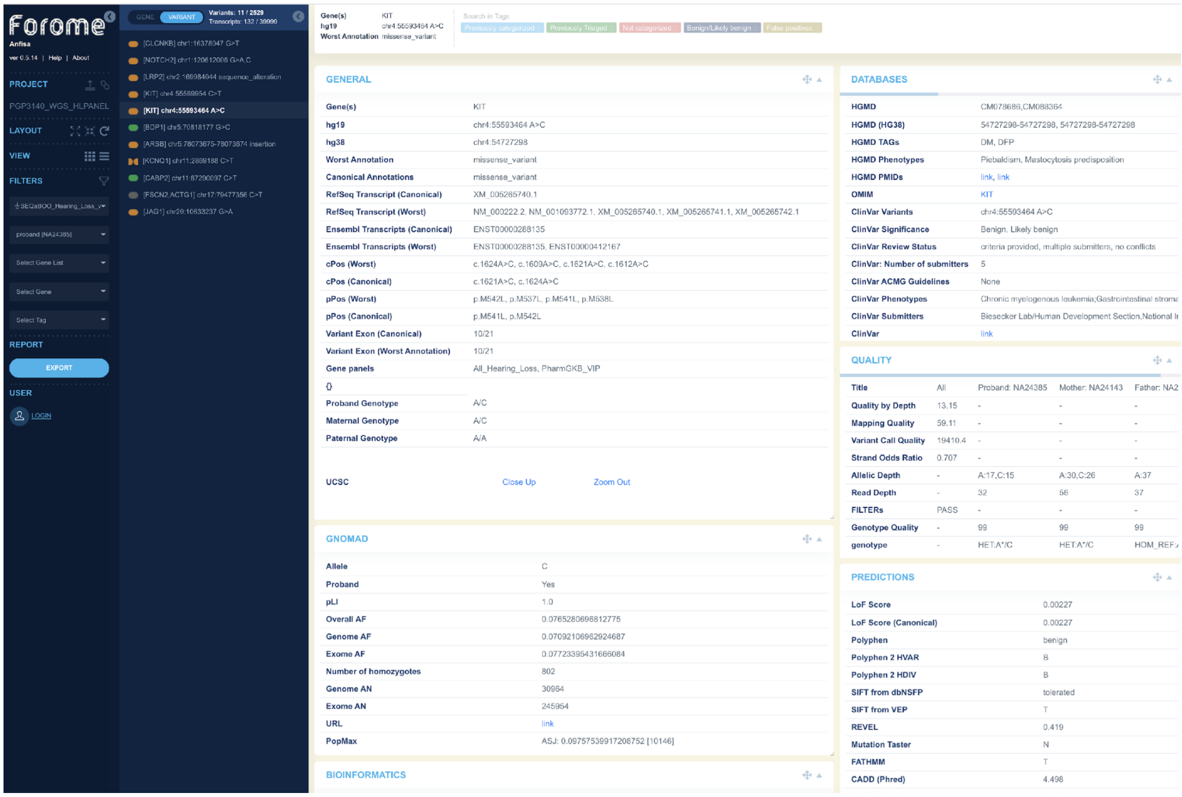


The next variant is chr5:71522350 G>C in *BDP1*. This one is more suspicious, because it is rare, though it is about 20 times more common in Ashkenazi Jews than in other populations and is not present in ClinVar or other databases. It is predicted to possibly cause a splice acceptor gain by the SpliceAI tool. It is a heterozygous variant in a gene associated with autosomal recessive hearing loss and inherited from the proband's mother, and as such is unlikely to cause hearing loss. This can be tagged for further review by the user.

The next *KCNQ1* variant chr11:2847958 C>T and the last variant in *JAG* should be categorized like the *KIT* one. The rest are extremely rare variants without clear clinical annotations. A user might want to look at the transcripts tab to categorize them. The transcripts that served as a base for selecting variants are shown in bold. If the user checks "Show Selection Only" checkbox, then all other transcripts become hidden.

#### Genetics first/Phenotype agnostic analysis

*Anfisa* includes two built-in decision trees which illustrate the Genetics-first approach: **BGM Red Button** (BGMRB) and **BGM Research** (BGMR). The BGMRB decision tree generates a subset of relatively high-quality variants, for which there are at least some reasons to suspect that they might be causal for a genetic defect. The BGMR decision tree is a superset of BGMRB that includes auxiliary variants which, while there is no known evidence of them being causative, there is no evidence to the contrary either, as well as variants with poorer call quality.

At the time of publication, a demo instance, based on *Anfisa* software version 0.6.12 and the Ashkenazi Trio high-confidence benchmarks VCF version 4.2 provided by NIST [56] is hosted at https://demo.forome.org/anfisa/app/dir.

We first review the *PGP3140_BGM_RedButton* data set created by application of BGMRB. The X-chromosome variants are not identified in the publicly available dataset; hence the X-linked inheritance mode is not applicable in this example. Therefore, we only review the autosomal dominant and recessive inheritance modes. Selecting the *Mendelian_Auto_Dom* filter yields no variant either. It is understandable given the nature of the dataset: autosomal dominant variants usually require either running dedicated *de-novo* callers or more than just 3 relatives included in the Variant Call Format file. In this dataset, however, we only have a simple trio.

Recessive analysis falls into two categories: homozygous and compound heterozygous. Selecting *Mendelian_Homozygous_Recessive* quick filter yields 6 variants in 5 genes, 5 missense and 1 in-frame insertion. All missense variants have mostly benign *in-silico* predictions and are not found in ClinVar. Detailed *in-silico* predictions can be examined in the **Transcripts** tab, where transcripts that served as the basis for inclusion of the variants in the list are shown in bold. By checking the "Show Selection Only" checkbox, the user can view only predictions relevant to the inclusion criteria. Selecting the *Mendelian_Compound_Het* filter displays a list of 7 compound heterozygous variants in 3 genes. They can be reviewed in the same way as the homozygous variants. Scrolling through the list with the open **Quality** tab provides detailed information about the alleles.

To get a better feel for how the filters are constructed, a user can click on the Conditions menu, click on the Filters button, then select Load and select one of the filters discussed above. Now the user can modify the filter by adjusting its options.

### Supplementary Note S9. Reporting chosen variants

#### Manual review and tagging of the variants

Once a user has chosen or built an appropriate filter for their analysis, they can manually review the selected variants. Each variant can be tagged with one or multiple tags. Most common tags are included in the default *Anfisa* package and users can add an unlimited number of their own tags. The user can also include a text note with each variant. Tags and notes are stored in the context of the primary datasets and are visible in all other datasets derived from the same source. An example of a tag can be: "**include in the report**".

#### Export options

*Anfisa* supports several ways of exporting selected variants. A selection can be based on filters, tags, or both.

To export selected variants as a Microsoft Excel Workbook, the user should first provide an export template. A template defines what properties of the variant are exported, in what order they are exported, and what colors/styles are used for specific columns. A sample template used for SEQaBOO project is included in the default package.

Variants can be also exported as a simple tab-delimited file. By default, only gene name and variant notations are included as two columns in the file. This option can be customized for a specific installation by modifying the *solutions.py* module.

Another option is to export variants in a JavaScript Object Notation (JSON) format that can be used for further machine processing.

Finally, the user can detach a derived dataset that can be made available to a different group of users as a primary dataset.

#### Supplementary Note 10. GitHub Repositories Structure

The *Forome* platform includes the following repositories:

***Anfisa*** (https://github.com/ForomePlatform/anfisa) is the main repository for the backend and built-in Graphical UI (GUI). It is developed in Python and contains implementation of most of the algorithms used for curation. Its subfolder https://github.com/ForomePlatform/anfisa/tree/master/app/config/files contains the source code for gene lists (panels) (extension *.lst) and built-in decision trees representing complex inclusion and exclusion criteria (extension *.pyt). Python module https://github.com/ForomePlatform/anfisa/blob/master/app/config/solutions.py contains implementations of built-in filters.

Functionality of the backend is exposed through REST API. The up to date description of the REST API is available at https://github.com/ForomePlatform/anfisa/blob/master/app/REST.txt

***Anfisa* Front End** (https://github.com/ForomePlatform/Anfisa-Front-End) is the repository for Vue.js based Front End. It communicates with the backend through the public REST API.

***Anfisa* Annotations** (https://github.com/ForomePlatform/Anfisa-Annotations) contains the annotation pipeline, which is a Java program.

**Variant Callers** (https://github.com/ForomePlatform/variant_callers) is the repository for standalone Python implementation of BGM callers including the Bayesian De-Novo caller and tools for creation of the library for the Bayesian De-Novo caller. This is a completely redeveloped implementation of the algorithm described in [47].
Additional repositories contain various utilities and projects at early stages of development.

### Code Sample. Built-in Curation Rules

For a full set of rules refer to:
https://github.com/ForomePlatform/anfisa/tree/master/app/config/files

#### Inclusion and Exclusion Criteria

##### Rare Variants for a Trio

```
#0.     Check sequencing quality
if Proband_GQ <= 19:
    return False
if Min_GQ <= 39:
    return False
if QD <= 4:
    return False
if FS >= 30:
    return False

#Always include De-Novo variants
if (Callers in {"BGM_BAYES_DE_NOVO"}):
    return True
if (Callers in {"RUFUS"}):
    return True
if (Callers in {"CNV"}):
    return True

#Exclude common variants
if gnomAD_AF_Genomes >= 0.01:
    return False
if gnomAD_AF_Exomes >= 0.01:
    return False
#Exclude known homozygous variants
if (gnomAD_Hom >= 1):
    return False
if (gnomAD_Hem >= 1):
    return False

#Exclude variants common for an ancestry group
if (gnomAD_PopMax_AN >= 2000 and gnomAD_PopMax_AF >= .05):
    return False

#Exclude non-coding
#   except those likely to alter splicing
if ((Most_Severe_Consequence in
            {
                "intergenic_variant",
                "intron_variant",
                "non_coding_transcript_exon_variant",
                "upstream_gene_variant",
                "downstream_gene_variant",
                "TF_binding_site_variant",
                "regulatory_region_variant",
                "5_prime_UTR_variant",
                "3_prime_UTR_variant",
                "splice_region_variant",
                "TFBS_ablation",
                "mature_miRNA_variant",
                "synonymous_variant"
            })
        and (splice_ai_dsmax <= 0.2)):
    return False

label("Comp-1")

# Inheritance Mode
if Inheritance_Mode() in {"Homozygous Recessive"}:
    return True
```

```
if Inheritance_Mode() in {"X-linked"}:
    return True

if Compound_Het(state="Comp-1") in {Proband}:
    return True
return False
```

#### Variants for BGM Analysis (Undiagnosed Patients)

```
#0.     Check sequencing quality
if Proband_GQ <= 19:
    return False

#Always include De-Novo variants
if (Callers in {"BGM_BAYES_DE_NOVO"}):
    return True
if (Callers in {"RUFUS"}):
    return True
if (Callers in {"CNV"}):
    return True
if (Variant_Class in {"CNV: deletion"}):
    return True

#Exclude common variants
if gnomAD_AF_Genomes >= 0.01:
    return False
if gnomAD_AF_Exomes >= 0.01:
    return False

#Exclude variants common for an ancestry group
if (gnomAD_PopMax_AN >= 2000 and gnomAD_PopMax_AF >= .05):
    return False

#Exclude very low impact variants
#   except those likely to alter splicing
if ((Most_Severe_Consequence in
            {
                "intron_variant",
                "intergenic_variant",
                "non_coding_transcript_exon_variant",
                "upstream_gene_variant",
                "downstream_gene_variant",
                "TF_binding_site_variant",
                "regulatory_region_variant"
            })
        and (splice_ai_dsmax <= 0.2)):
    return False

label("Comp-1")

# Inheritance Mode
if Inheritance_Mode() in {"Homozygous Recessive"}:
    return True

if Inheritance_Mode() in {"X-linked"}:
    return True

if Inheritance_Mode() in {"Autosomal Dominant"}:
    return True

if Compound_Het(state="Comp-1") in {Proband}:
    return True

return False
```

#### Hearing Loss variants

```
#0.     Check sequencing quality
if Proband_GQ <= 19:
    return False
if FS > 30:
    return False
if QD < 4:
    return False
#Exclude  variants not detected in the family
if Num_Samples < 1:
    return False
#Exclude  variants not in hearing loss panel
if Panels not in {All_Hearing_Loss}:
    return False
#Include Present in HGMD as "DM"
if HGMD_Tags in {"DM"}:
    return True
# Exclude common variants AF> 5%
if gnomAD_AF >= 0.05:
    return False
#Exclude variants farther then 5pb from intronic/exonic border
if (not Region in {"exon"}) and Dist_from_Exon >= 6:
    return False
#2.a.  Include if present in ClinVar as: Path, Likely Path, VUS
# (worst annotation, unless annotated benign by trusted submitter')
if (Clinvar_Benign in {"False"} and
        Clinvar_Trusted_Benign in {"False", "No data"}):
    return True
# 2.b. Include All de novo variants
if (Callers in {"BGM_BAYES_DE_NOVO"}):
    return True
if (Callers in {"RUFUS"}):
    return True
if (Callers in {"CNV"}):
    return True
# 2.c. Include all potential LOF variants
#       (stop-codon, frameshift, canonical splice site).
if (Most_Severe_Consequence in {
            'transcript_ablation',
            'splice_acceptor_variant',
            'splice_donor_variant',
            'stop_gained',
            'frameshift_variant',
            'stop_lost',
            'start_lost'
        }):
    return True
# 3.a. Leave only:
#   "Missense", "synonymous" and "splice region" variants
if (Most_Severe_Consequence not in {
        "inframe_insertion",
        "inframe_deletion",
        "missense_variant",
        "protein_altering_variant",
        "splice_region_variant",
        "synonymous_variant",
        "stop_retained_variant",
        "coding_sequence_variant"
        }):
    return False
#3.    Include: AF < 0.0007 (GnomAD Overall)
#  And: PopMax < 0.01
#       (minimum 2000 alleles total in ancestral group)')
if (gnomAD_AF <= .0007 and
        (gnomAD_PopMax_AN <= 2000 or gnomAD_PopMax_AF <= .01)):
    return True

return False
```

#### Variants with Damaging Predictions

```
if (Most_Severe_Consequence in {
            'transcript_ablation',
            'splice_acceptor_variant',
            'splice_donor_variant',
            'stop_gained',
            'frameshift_variant',
            'CNV: deletion',
            'start_lost'
        }):
    return True

if (Clinvar_stars in {'2', '3', '4'} and
        ClinVar_Significance in {
            'Likely pathogenic',
            'Pathogenic',
            'Pathogenic, Affects',
            'Pathogenic, other',
            'Pathogenic, protective',
            'Pathogenic, association, protective',
            'Pathogenic, drug response',
            'Pathogenic, other, risk factor',
            'Pathogenic, risk factor',
            'Pathogenic/Likely pathogenic',
            'Pathogenic/Likely pathogenic, drug response',
            'Pathogenic/Likely pathogenic, risk factor',
            'Pathogenic/Likely pathogenic, other',
            'Likely pathogenic, drug response',
            'Likely pathogenic, risk factor',
            'Likely pathogenic, other',
            'Likely pathogenic, association'
        }):
    return True

if (Clinvar_stars in {'2', '3', '4'} and
        ClinVar_Significance in {
            'Benign',
            'Benign, association',
            'Benign, drug response',
            'Benign, other','Benign, risk factor',
            'Benign/Likely benign',
            'Benign/Likely benign, Affects',
            'Benign/Likely benign, association',
            'Benign/Likely benign, drug response',
            'Benign/Likely benign, drug response, risk factor',
            'Benign/Likely benign, other','Benign/Likely benign, protective',
            'Benign/Likely benign, protective, risk factor',
            'Benign/Likely benign, risk factor'
        }):
    return False

if splice_ai_dsmax > 0.5:
    return True
if Polyphen_2_HVAR in {"P", "D"}:
    return True
if Polyphen_2_HDIV in {"B"}:
    return False
if SIFT in {"deleterious", "deleterious_low_confidence"}:
    return True
if SIFT in {"tolerated", "tolerated_low_confidence"}:
    return False
if Polyphen_2_HDIV in {"P", "D"}:
    return True
if Polyphen_2_HVAR in {"B"}:
    return False

return False
```

#### Simple Filters

Most of the filters below are supposed to be applied to workspaces created by applying “Variants for BGM Analysis” and Exclusion Criteria or a similar curation rule.

##### Common for all Mendelian Filters

- **FT in {PASS}**
- **Proband_GQ >= 50**
- **Min_GQ >= 40 # Minimum GQ for all family members**
- **QD >= 4**
- **FS <= 30**
- **Transctipt_consequence in {**
  - **"CNV: deletion",**
  - **transcript_ablation,**
  - **splice_acceptor_variant,**
  - **splice_donor_variant,**
  - **stop_gained, frameshift_variant,**
  - **inframe_insertion,**
  - **inframe_deletion,**
  - **missense_variant,**
  - **protein_altering_variant,**
  - **incomplete_terminal_codon_variant,**
  - **synonymous_variant,**
  - **splice_region_variant,**
  - **coding_sequence_variant**
- **}**
- **Transcript_biotype in {protein_coding}**
- **Transcript_source in {Ensembl}**

##### Mendelian Homozygous Recessive

Common for all Mendelian filters plus:

- **Inheritance_Mode() in {"Homozygous Recessive"}**

##### Mendelian Compound Heterozygous

Common for all Mendelian filters plus:

- **Compound_Het(approx = "transcript") in {Proband}**

##### Mendelian Autosomal Dominant

Common for all Mendelian filters plus:

- **Inheritance_Mode() in {" Autosomal Dominant"}**

##### BGM De-Novo

- **Transctipt_consequence in {**
    - **"CNV: deletion",**
    - **transcript_ablation,**
    - **splice_acceptor_variant,**
    - **splice_donor_variant,**
    - **stop_gained, frameshift_variant,**
    - **inframe_insertion,**
    - **inframe_deletion,**
    - **missense_variant,**
    - **protein_altering_variant,**
    - **incomplete_terminal_codon_variant,**
    - **synonymous_variant,**
    - **splice_region_variant,**
    - **coding_sequence_variant**
- **}**
- **Callers in {BGM_BAYES_DE_NOVO, RUFUS}**

##### Impact Splicing

- **FT in {PASS}**
- **Proband_GQ >= 50**
- **Min_GQ >= 40 # Minimum GQ for all family members**
- **QD >= 4**
- **FS <= 30**
- **splice_ai_dsmax > 0.2**

#### Supplementary Table ST1. Examples of Available Variant Curation Platforms

| **Name** | **Country** | **References** |
|---|---|---|
| Genuity Science (Wuxi outside of China) | US | |
| Illumia TruSight (incl BlueBee acq) | US | https://www.illumina.com/informatics/biological-interpretation/variant-analysis/rare-variants.html |
| Broad Institute Seqr (US) | US | |
| Sophia Genetics (w/acquired Interactive Biosoftware) | CH | |
| Agilent (w/acquired acquiring Alissa Interpret) (US) | US | |
| Breakthrough Genomics Eliter | US | https://btgenomics.com/#technology<br>https://enliter.btgenomics.org/user/login |
| Invitae (US) incl Genosity & Sherloc | US | https://www.genosity.com/software/integrated-genomic-toolkit/<br>https://www.invitae.com/en/variant-classification/ |

| | | |
|---|---|---|
| Opko Health GeneDX | | https://www.genedx.com/wp-content/uploads/2020/08/40150_Clinical-Genomics-Overview-Brochure-FINAL-07.2020.pdf |
| Congenica | UK | |
| Emedgene | IL | |
| Fabric Genomics | US | https://fabricgenomics.com/fabric-gem/ |
| Saphetor Varsome | CH | https://saphetor.com/<br>https://varsome.com/ |
| GeneYX | IL | https://geneyx.com/geneyxanalysis/ |
| Nostos Genomics | DE | https://www.nostos-genomics.com/#product |
| Genoox, Genoox Franklin | US | https://franklin.genoox.com/clinical-db/home<br>https://www.genoox.com/bioinformatics-technology/genomic-tools/ |
| GenomCore<br>(+ Made of Genes - e2e clinical services) | ES | https://genomcore.com/en/technology-stack/ |
| PierianDx (+Knome + Tute Genomics) | US | https://www.pieriandx.com/clinical-genomics-software-for-next-generation-sequencing |
| Ranomics | US | https://www.ranomics.com/solutions |
| Qlucore | SE | https://www.qlucore.com/products |
| Engenome (eVai Interpreter) | IT | https://www.engenome.com/product/ |

#### Supplementary Table ST2. Color and Shape codes used for variants visualization

Algorithm for sequence variant's classification used in *Anfisa* for color and shape coded variant's labels.

<table>
<tr><td></td><td>HGMD DM</td><td>HGMD DM?</td><td>HGMD other or absent</td></tr>
<tr><td>ClinVar pathogenic</td><td>Consensus pathogenic</td><td>Consensus pathogenic</td><td>Consensus pathogenic</td></tr>
<tr><td>ClinVar benign</td><td>Consensus benign</td><td>Consensus benign</td><td>Consensus benign</td></tr>
<tr><td>ClinVar other or absent</td><td>Consensus pathogenic</td><td>Consensus pathogenic</td><td>In-Silico Predictions
<table>
<tr><td></td><td>Worst benign</td><td>Worst possibly damaging</td><td>Worst damaging</td></tr>
<tr><td>Best Benign</td><td>Consensus benign</td><td>Uncertain</td><td>Uncertain</td></tr>
<tr><td>Best possibly damaging</td><td>X</td><td>Uncertain</td><td>Uncertain</td></tr>
<tr><td>Best damaging</td><td>X</td><td>X</td><td>Consensus pathogenic</td></tr>
</table>
No in-silico predictions: absent</td></tr>
</table>

#### Supplementary Table ST3. Ratios of Damaging to Benign Variants for Sepsis Patients with and without PF

| | | Complement System | | Coagulation System | |
|---|---|---|---|---|---|
| | | PF Cohort | Control Cohort | PF Cohort | Control Cohort |
| PolyPhen 2 HDIV | Damaging | 61 | 28 | 32 | 16 |
| | Benign | 61 | 41 | 48 | 23 |
| | **Ratio** | **1.00** | **0.68** | **0.67** | **0.70** |
| Polyphen 2 HVAR | Damaging | 49 | 20 | 25 | 12 |
| | Benign | 77 | 55 | 56 | 28 |
| | **Ratio** | **0.64** | **0.36** | **0.45** | **0.43** |
| SIFT | Damaging | 87 | 53 | 41 | 20 |
| | Benign | 77 | 53 | 49 | 29 |
| | **Ratio** | **1.13** | **1.00** | **0.84** | **0.69** |
| FATHMM | Damaging | 42 | 19 | 54 | 32 |
| | Benign | 114 | 81 | 37 | 17 |
| | **Ratio** | **0.37** | **0.23** | **1.46** | **1.89** |
